# A Human Glomerular Disease Atlas defines the APOL1-JAK-STAT feed forward loop in focal segmental glomerulosclerosis

**DOI:** 10.1101/2025.09.12.25335572

**Authors:** Phillip J. McCown, Charlotte Boys, Somenath Datta, Viji Nair, Edgar Otto, Damian Fermin, John Hartman, Francesca Annese, Fadhl Alakwaa, Khun Zaw Latt, Cathy Smith, Bradley Godfrey, Maria Larkina, Alison E. Ashley-Koch, Melanie E. Garrett, Savannah Moore, Karen Soldano, Mathew Alaba, Michelle T. McNulty, Dongwon Lee, Matthew G. Sampson, Laura H. Mariani, Abhijit S. Naik, Robin Fallegger, Sean Eddy, Julio Saez-Rodriguez, Opeyemi A. Olabisi, Matthias Kretzler, the Nephrotic Syndrome Study Network (NEPTUNE)

## Abstract

There are few approved treatments for glomerular diseases of the kidney. To map underlying transcriptional programs in the kidney driving rare glomerular diseases, single-nucleus RNA sequencing (snRNAseq) on kidney biopsies (N=120) from the Nephrotic Syndrome Study Network were integrated with snRNAseq and single-cell sequencing (scRNAseq) of reference kidney tissue (N=50) to create the Omnibus of CElls And Nuclei (OCEAN). Unsupervised analysis of multi-cellular programs identified that JAK-STAT pathway activity was associated with clinical measures of disease severity. JAK-STAT pathway activity was strongly correlated with apolipoprotein1 (APOL1) mRNA transcript expression and the high risk APOL1 variant genotype, a major risk factor for focal segmental glomerulosclerosis. These findings were confirmed in an independent study of Black participants where loss of APOL1 function decreased JAK-STAT pathway activation in *ex vivo* models of patient-derived podocytes. The findings presented are consistent with a feed forward loop regulating the JAK-STAT-APOL1 driven tissue damage, providing mechanistic support for the JUSTICE Phase II trial targeting JAK activation in APOL1-mediated kidney disease.

## Introduction

Kidney diseases are a global health threat^1^. Among these, primary glomerular diseases, which include focal segmental glomerulosclerosis (FSGS), minimal change disease (MCD), and IgA nephropathy (IgAN), though individually rare, are associated with debilitating symptoms and contribute substantially to the kidney failure pandemic^2^. Glomerular diseases are often characterized by nephrotic syndrome (NS), hallmarked by proteinuria, hypoalbuminemia, hyperlipidemia, and edema. Diagnosis is based on histological features related to patterns of injury in kidney biopsies that determine treatment course^3^. The only FDA-approved medical treatments are prednisone and adrenocorticotrophic hormone, which do not prevent progression to kidney failure in most patients and cause severe side effects. Guideline-based^3^ clinical care includes off-label use of second line immunosuppressive medications. The variability in response to these immunosuppressives, including steroids, adds to the disease burden. While many therapies are currently under development^4,5^ none have been approved specifically for FSGS and, until recently^6–8^, no therapies were approved specifically for any glomerular disease.

The heterogeneity of etiology and disease progression, and paucity of treatment options, highlight the essential prerequisite of a comprehensive knowledge environment defining the clinical, structural and molecular determinants of glomerular diseases to develop and implement effective treatments. To address this challenge, the Nephrotic Syndrome Study Network (NEPTUNE) has prospectively enrolled and followed participants with glomerular diseases over the last 17 years^9^. The NEPTUNE study has expanded our understanding of molecular mechanisms in FSGS^10,11^ and APOL1-mediated kidney disease (AMKD)^12,13^.

The presence of select apolipoprotein L1 (APOL1) single nucleotide polymorphisms (SNPs) in people of African ancestry is known to contribute to increased risk of developing AMKD, with worse outcomes for those with FSGS^14–17^. Increased expression of APOL1 variants (G1 and G2) and not the reference APOL1 allele (G0) in podocytes, specialized cells in the glomerular filter compartment of kidneys, caused dose-dependent injury^18,19^ and FSGS^20^. Meanwhile, a specific APOL1 variant (N264K) that inactivates APOL1 has been identified to exhibit kidney protective effects^19,21,22^. Despite the causal role of APOL1 in kidney injury, only 20% of carriers of the high-risk APOL1 genotype are estimated to develop AMKD^14,23^. The basis of this incomplete penetrance is unknown.

The advent of single nucleus RNA sequencing (snRNAseq) and single cell RNA sequencing (scRNAseq) technologies offers an unprecedented opportunity to understand molecular mechanisms of disease and identify novel cell-states and niches. Multi-institutional research consortia, such as the Human Cell Atlas^24,25^ and the Kidney Precision Medicine Project^26,27^ have provided key insights into human health and disease at a cellular-level. NEPTUNE now deploys such mechanistic disease definitions to develop and test targeted therapies^28^.

In this study, kidney biopsies contributed by NEPTUNE participants and kidney donor samples from the Human Kidney Transplant Transcriptomic Atlas (HKTTA), as reference tissue, were used to generate and integrate snRNAseq and scRNAseq profiles to create the <u>O</u>mnibus of <u>CE</u>lls <u>A</u>nd <u>N</u>uclei (OCEAN) dataset (https://neptune-ocean.cxg.miktmc.org/). An unsupervised analysis of the OCEAN data identified multi-cellular programs underlying glomerular disease heterogeneity and enabled cell-type specific explorations in FSGS leading to the characterization of JAK-STAT signaling and APOL1 interactions. Transcriptomic, genetic and proteomic analyses followed by validation using human podocytes derived from induced pluripotent stem cells (IPSCs) enabled detailed investigations into the molecular interactions in glomerular diseases, particularly in the context of the APOL1 risk allele (**Fig. 1**). Importantly, loss of APOL1 function was discovered to be associated with attenuation of JAK-STAT activation, consistent with a feed-forward regulatory loop between APOL1 and JAK-STAT. These findings further support the JUSTICE trial^29^, part of the NEPTUNE Match precision medicine trial platform^28^, evaluating JAK inhibitors in people with APOL1 risk variants. This study is illustrative of the power of the OCEAN dataset to drive hypothesis generation for further mechanistic studies and clinical trials in rare glomerular diseases.

**Figure 1.**
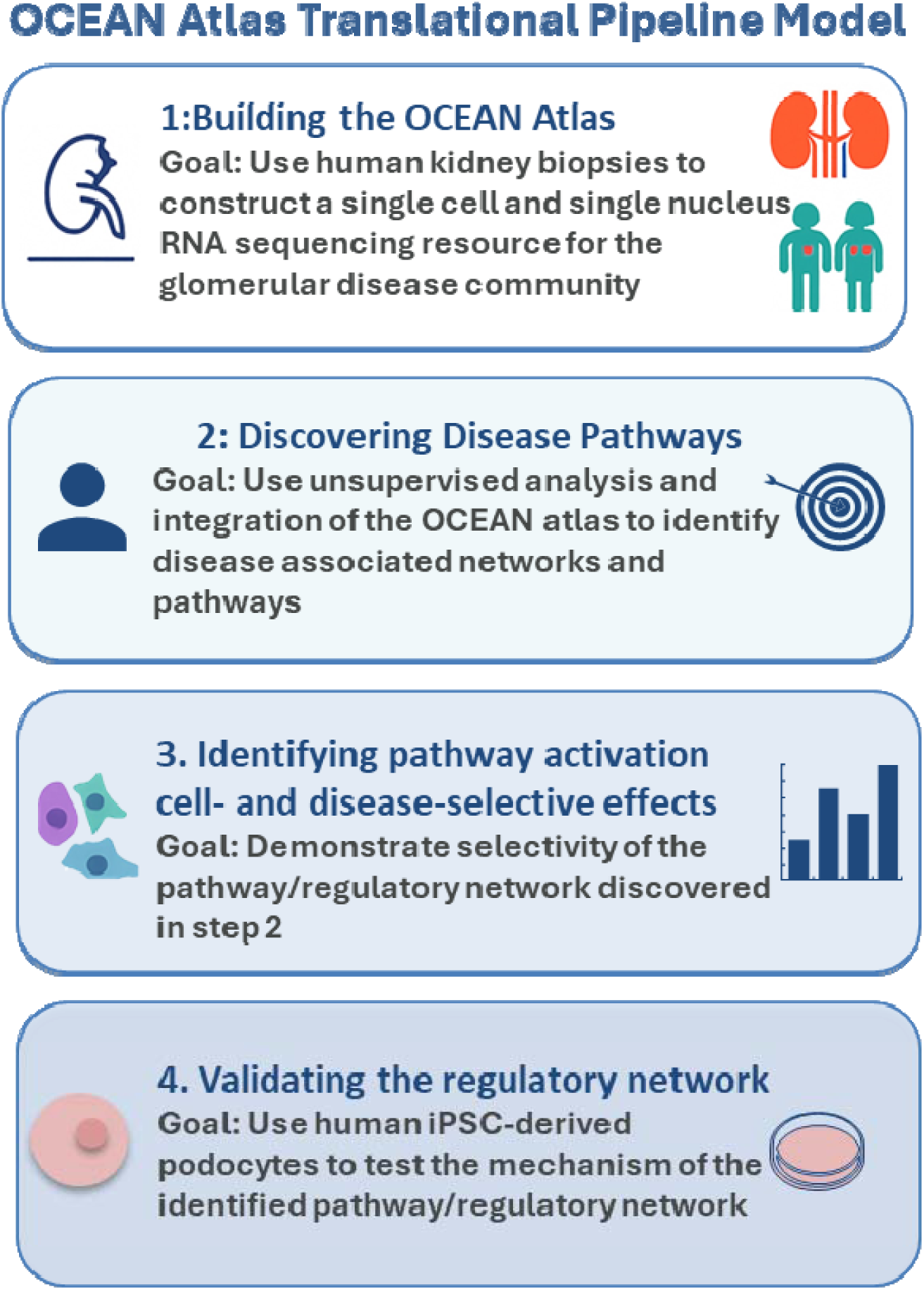
Overview of study’s analytical approach. This figure presents the high-level analysi framework for “A Human Glomerular Disease Atlas defines the APOL1-JAK-STAT feed forward loop in focal segmental glomerulosclerosis”.

## Results

### Building the OCEAN - representing a comprehensive collection of glomerular disease kidney cell types

We derived the OCEAN dataset by sequencing kidney tissues of 120 participants obtained at the time of enrollment (baseline diagnostic biopsy) into NEPTUNE (**Table 1**) and stored in RNAlater for up to 10 years. The number of nuclei captured was not affected by time in storage. Reference data in OCEAN comprise 49 kidney tissue profiles (scRNAseq and snRNAseq) from HKTTA (transplanted kidneys, **Table 1**) obtained from living (LD) and deceased donor kidney biopsies. NEPTUNE recruited participants prior to biopsy based on clinical profile, capturing a diverse group of participants as subsequently defined by descriptive histopathology adjudication. Participants were predominantly diagnosed with FSGS (N = 33), MCD (N = 24) or proteinuric IgAN (N = 29)^9^. The dataset also includes biopsies from 34 participants with other proteinuric diseases or those pending histological adjudication. snRNAseq and/or scRNAseq profiles were integrated into an atlas of 1,047,781 nuclei and cells (**Fig. 2a**) that formed 17 major cell clusters (**Fig.2b**) and 28 discrete clusters (**SFig. S1**), representing all the nephron cell types from both glomerular and tubulointerstitial compartments of the kidney. The number of nuclei contributed were similar for FSGS, MCD, and IgAN, and cell numbers were proportionate to the number of participants in each group (**Fig. 2c**, **Table 1**). Clusters were preserved across all major diagnoses and transplant sample types (**SFig. S1b-h**). Since biopsies are typically obtained from the kidney cortex, as expected, there were more cells in OCEAN from the tubular compartment, with the most from the proximal tubule (PT) and thick ascending limb (TAL) (**Fig 2d**). The distribution of resident kidney cells across the main groups (LD, FSGS, MCD and IgAN) were similar with all groups contributing cells to each cell type with a slightly increased contribution of PT cells from MCD. We also observed fewer cells in the POD cluster from FSGS and IgAN, as expected, since podocyte loss is common in these conditions where glomeruli are scarred (**Fig 2e**).

**Figure 2.**
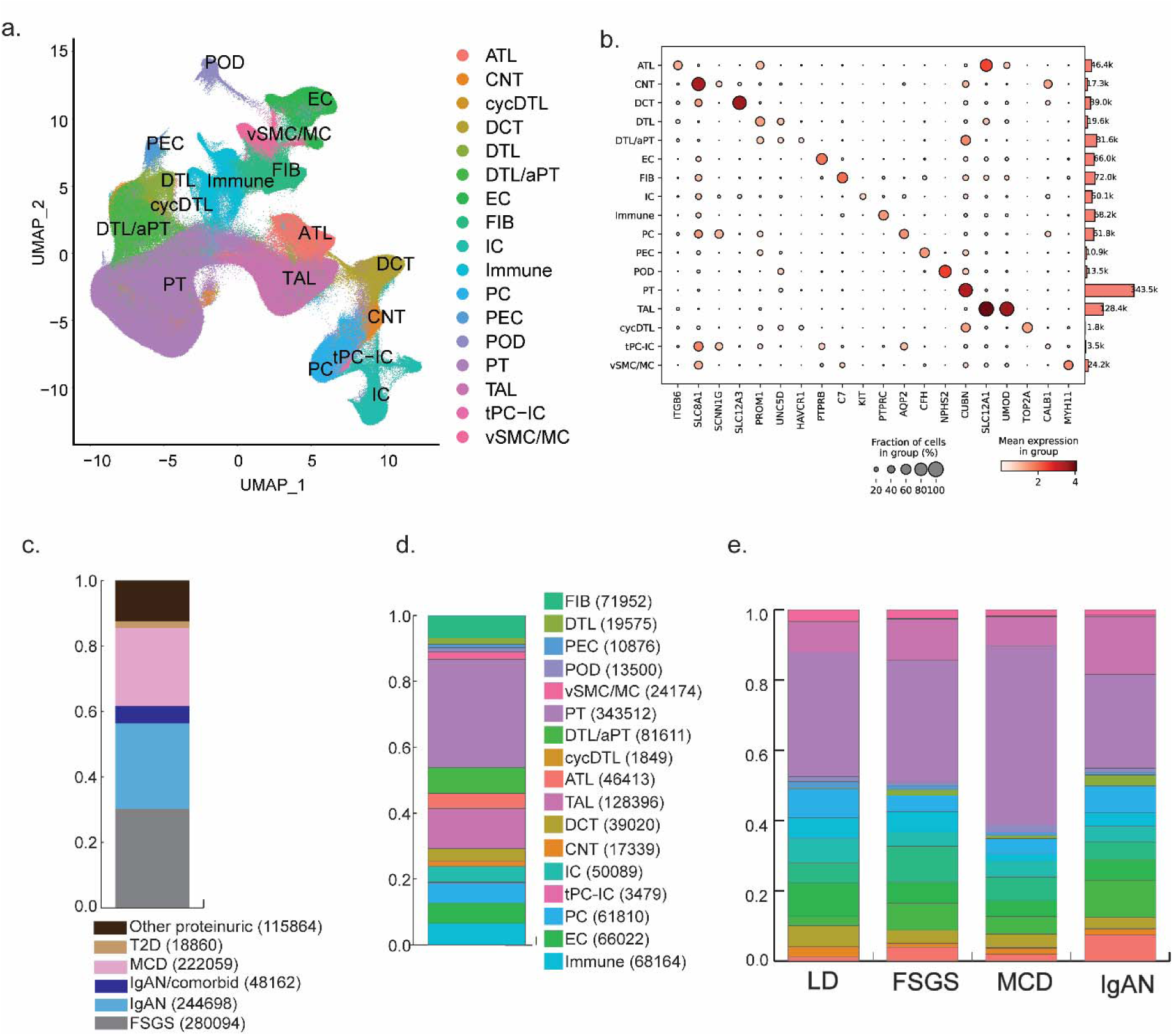
OCEAN composition and distribution. **a.** UMAP of 1,047,781 nuclei and cells within OCEAN revealed all nephron and several non-nephron cell types. **b.** The cell type that represented the majority membership of each Seurat cluster was inferred based on cell type markers and common transcriptomic signatures associated with cells seen in KPMP and PREMIERE. **c.** Proportional distribution of the nuclei of OCEAN based on participants with FSGS (n=33), MCD (n=24), IgAN (n=29), IgAN comorbid with a different kidney disease (n=4), T2D (n=2), LD (n=11), or other proteinuric diseases (n=28). **d.** Overall proportional composition of OCEAN containing both snRNAseq and scRNAseq datasets. **e.** Proportional composition of nuclei within OCEAN across LD, FSGS, MCD, and IgAN. Abbreviations: KPMP-Kidney Precision Medicine Project; PREMIERE - PREcision Medicine through IntErrogation of Rna in the kidnEy. FIB = fibroblast, PEC = parietal epithelium, POD = podocyte, vSMC/MC = vascular smooth muscle and mesangium, PT = proximal tubule, DTL = descending thin limb, aPT = adaptive proximal tubule, cycDTL = cycling descending thin limb, ATL = ascending thin limb, TAL = thick ascending limb, DCT = distal convoluted tubule, CNT = connecting tubule, IC = intercalated cell, tPC-IC = transitioning principal/intercalated cell, PC = principal cell, EC = endothelial cell, Immune = immune cells. FSGS-focal segmental glomerulosclerosis; MCD-minimal change disease; IgAN - IgA nephropathy; LD – living donor.

**Table 1.**
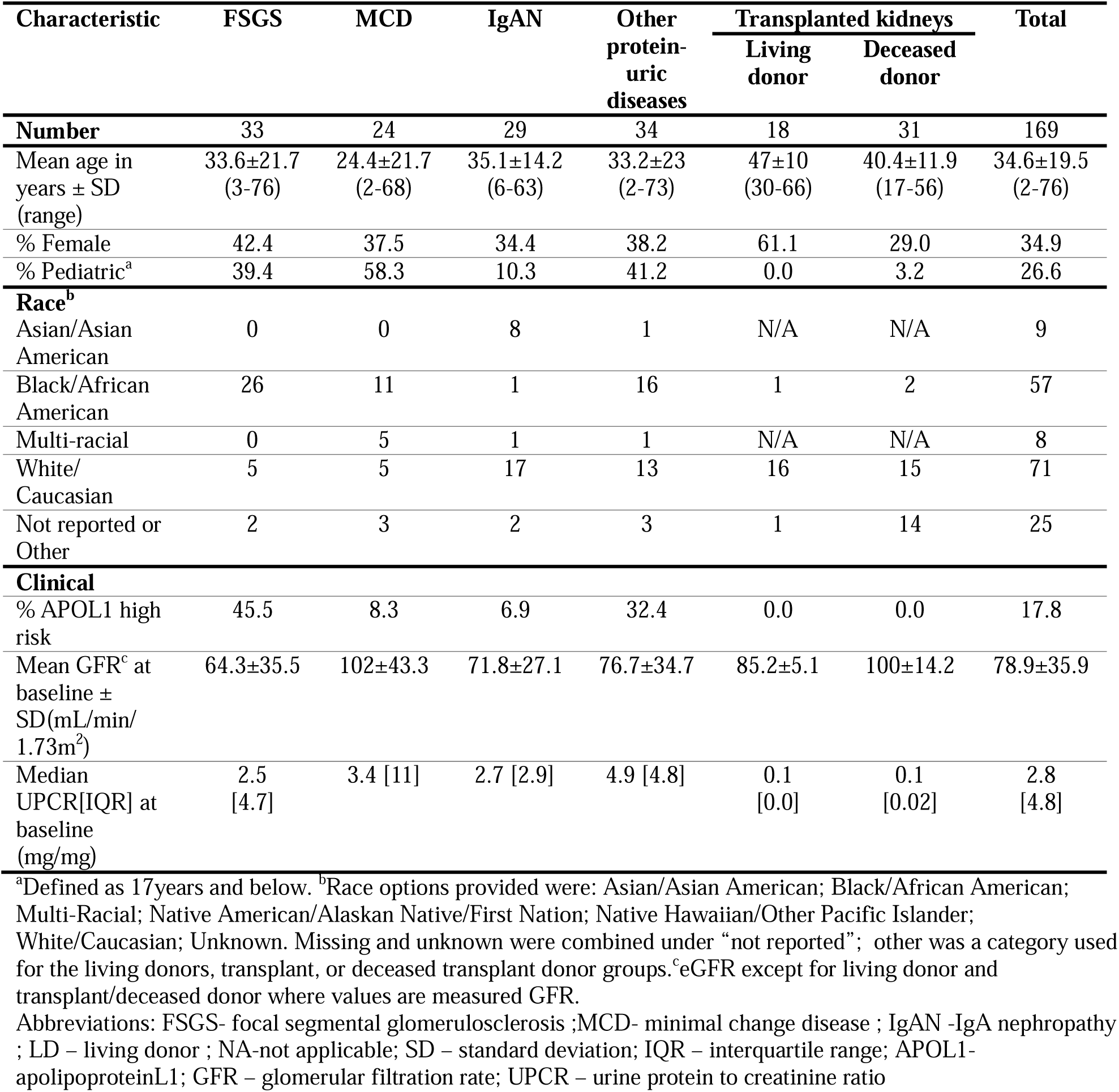
Clinical and demographic characteristics of OCEAN participants.

| Characteristic | FSGS | MCD | IgAN | Other protein-uric diseases | Transplanted kidneys |  | Total |
| --- | --- | --- | --- | --- | --- | --- | --- |
|  |  |  |  |  | Living donor | Deceased donor |  |
| **Number** | 33 | 24 | 29 | 34 | 18 | 31 | 169 |
| Mean age in years ± SD (range) | 33.6±21.7 (3-76) | 24.4±21.7 (2-68) | 35.1±14.2 (6-63) | 33.2±23 (2-73) | 47±10 (30-66) | 40.4±11.9 (17-56) | 34.6±19.5 (2-76) |
| % Female | 42.4 | 37.5 | 34.4 | 38.2 | 61.1 | 29.0 | 34.9 |
| % Pediatric<sup>a</sup> | 39.4 | 58.3 | 10.3 | 41.2 | 0.0 | 3.2 | 26.6 |
| **Race<sup>b</sup>** |  |  |  |  |  |  |  |
| Asian/Asian American | 0 | 0 | 8 | 1 | N/A | N/A | 9 |
| Black/African American | 26 | 11 | 1 | 16 | 1 | 2 | 57 |
| Multi-racial | 0 | 5 | 1 | 1 | N/A | N/A | 8 |
| White/Caucasian | 5 | 5 | 17 | 13 | 16 | 15 | 71 |
| Not reported or Other | 2 | 3 | 2 | 3 | 1 | 14 | 25 |
| **Clinical** |  |  |  |  |  |  |  |
| % APOL1 high risk | 45.5 | 8.3 | 6.9 | 32.4 | 0.0 | 0.0 | 17.8 |
| Mean GFR<sup>c</sup> at baseline ± SD(mL/min/1.73m<sup>2</sup>) | 64.3±35.5 | 102±43.3 | 71.8±27.1 | 76.7±34.7 | 85.2±5.1 | 100±14.2 | 78.9±35.9 |
| Median UPCr[IQR] at baseline (mg/mg) | 2.5 [4.7] | 3.4 [11] | 2.7 [2.9] | 4.9 [4.8] | 0.1 [0.0] | 0.1 [0.02] | 2.8 [4.8] |

<sup>a</sup>Defined as 17years and below. <sup>b</sup>Race options provided were: Asian/Asian American; Black/African American; Multi-Racial; Native American/Alaskan Native/First Nation; Native Hawaiian/Other Pacific Islander; White/Caucasian; Unknown. Missing and unknown were combined under “not reported”; other was a category used for the living donors, transplant, or deceased transplant donor groups. <sup>c</sup>eGFR except for living donor and transplant/deceased donor where values are measured GFR.
Abbreviations: FSGS- focal segmental glomerulosclerosis ;MCD- minimal change disease ; IgAN -IgA nephropathy ; LD – living donor ; NA-not applicable; SD – standard deviation; IQR – interquartile range; APOL1- apolipoproteinL1; GFR – glomerular filtration rate; UPCr – urine protein to creatinine ratio

### Discovery of disease associated pathways - Unsupervised analysis of OCEAN transcriptomic data provides molecular context to patient heterogeneity in glomerular diseases

Multicellular factor analysis (MCFA)^30^, a variance decomposition method which can be conceptualized as an extension of principal component analysis (PCA) across multiple data ‘views’, was used to quantify inter-patient variation in the molecular data. Data views included gene expression profiles aggregated at the major cell type level (**STable S1**) and inferred ligand-receptor interactions between cell types. MCFA identified a set of latent factors, each describing a major axis of variation across participants. Patient samples received a score for each factor, which were characterized by gene weights within each cell type and ligand-receptor weights for sender-receiver cell pairs^31^ (**Fig. 3a**). This analysis was carried out on the subset of snRNAseq samples within OCEAN profiled on the 10x platform comprising 97 NEPTUNE and 11 reference samples across 850,703 sequenced nuclei (**Fig. 3b**).

**Figure 3.**
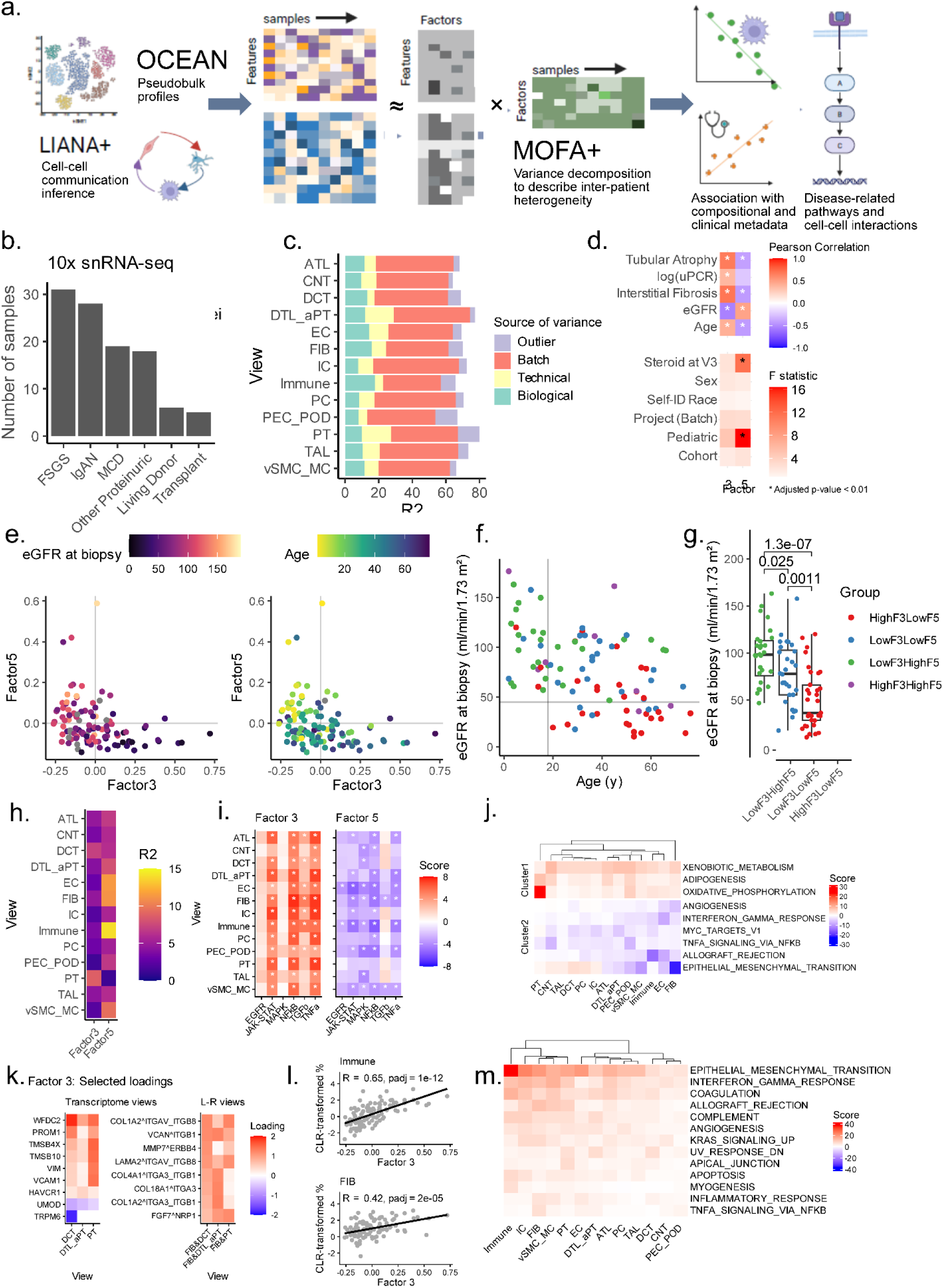
Unsupervised exploratory analysis of OCEAN reveals molecular details of kidney function decline **a.** Schematic of the unsupervised analysis using Multicellular Factor Analysis (MCFA)^30^ to define the overall Factor space and characterize it in terms of pathways^32^ and cell-cell interactions^31^. **b.** Overview of samples included in the unsupervised analysis. **c.** Total variance explained (R2) across all factors for each of the cell-type views, categorized by source of variance. **d.** Post-hoc association with clinical metadata of interest. Pearson correlation coefficient shown for continuous metadata (top), ANOVA F-statistic shown for categorical metadata (bottom). *-FDR-adjusted p-value < 0.01**. e.** A two-dimensional map of MCFA Factors 3 and 5 puts OCEAN biopsies on a spectrum of eGFR (ml/min/1.73m²) and age (years) at biopsy. **f-g.** Age and eGFR distribution of biopsies by Factor 3-Factor 5 stratification group. Two-tailed t-test. **h.** Variance explained per cell-type view for Factor 3 and Factor 5. **i.** PROGENy pathway activity enrichment on gene loadings of Factor 3 and Factor 5. *-FDR-adjusted p-value < 0.01. **j.** Hallmark gene set enrichment on Factor 5 loadings. Score indicates -log10(FDR.p.value) on terms enriched among positive loadings, and log10(FDR.p.value) on terms enriched among negative loadings. **k.** Selected loadings of Factor 3 in transcriptomic views (left) and ligand-receptor views (right). **l.** Post-hoc association (Pearson correlation) between Factor 3 score and center log-ratio transformed cell type proportions of cells annotated Immune and FIB. **m.** Hallmark gene set enrichment on Factor 3 loadings. Score indicates -log10(FDR.p.value) on terms enriched among top 200 positive loadings, and log10(FDR.p.value) on terms enriched among top 200 negative loadings.

MCFA with 20 factors explained 63–75% of inter-sample variance in gene expression per cell type (**Fig. 3c**). We categorized thesefactors into four groups: 1) batch factors, which primarily distinguished samples processed in different experimental batches, likely due to technical variation in sequencing or sample handling; 2) outlier factors, which captured variability driven by a single sample deviating from the cohort; 3) technical factors, which separated samples based on method of tissue retrieval, or the relative proportion of medullary cells profiled; and 4) biological factors, which accounted for 8–18% of variance and reflected disease-related gene expression changes. The two biological factors, Factor 3 and Factor 5, were significantly associated with kidney function at the time of biopsy as measured by eGFR (Pearson r = −0.55, p-adj = 1.25e-07 and r = 0.47, p-adj = 1.28e-05 respectively), as well as other clinical covariates (**Fig.3d**). Thus, the unsupervised exploration distinguished biological from technical variability and enabled further investigations into variations related to disease.

### Discovery of disease associated pathways - Two axes of variation reveal a “cell state map” of kidney function decline in glomerular disease

To explore the biological and clinical relevance of these factors, we examined their relationship to participant demographics and kidney function. Factor 3 and Factor 5 scores were orthogonal, representing two distinct axes of variation. A two-dimensional map, with participant coordinates based on their Factor 3 and Factor 5 scores, helped visualize heterogeneity in disease severity (**Fig. 3e**). Together, they define a molecular spectrum of kidney function in glomerular failure, ranging from younger individuals with preserved kidney function to older individuals with progressive decline in eGFR (**Fig. 3e**). We stratified the two-dimensional participant map into four quadrants: HighF3LowF5, LowF3LowF5, LowF3HighF5, and HighF3HighF5 groups, where ‘High’ corresponds to a factor score > 0 and ‘Low’ to a factor score < 0. Young participants with higher eGFR (age < 18 years, eGFR ≥ 45 ml/min/1.73m²) mainly clustered in the LowF3HighF5 group (17 out of 28). In contrast, adults with low eGFR (age ≥ 18 years, eGFR < 45 ml/min/1.73m²) were predominantly (17 out of 24) grouped in the HighF3LowF5 quadrant (**Fig. 3f**). Overall, eGFR decreased significantly and progressively between the LowF3HighF5 group (best kidney function) to LowF3LowF5 (middle kidney function) (p = 0.025), and from LowF3LowF5 (middle kidney function) to HighF3LowF5 (worst kidney function) (p = 0.0011; **Fig. 3g**). This progressive decline in kidney function between the three quadrants held when constricting to the adult cohort (age ≥ 18 years), where there was significantly lower eGFR in the HighF3LowF5 group compared to the LowF3HighF5 group (p = 1.1e-05), but not the pediatric cohort (age < 18 years), where there was no statistically significant difference (p = 0.32) (**SFig. S2a**). These findings suggest that Factor 3 and Factor 5 provide a molecular readout of the cell states of the kidney at different levels of eGFR and may be useful for stratifying adult patients along a trajectory of kidney function decline.

### Discovery of disease associated pathways - Multicellular program of kidney function decline: decreased metabolic activity and the establishment of a profibrotic state

Factor 5, which positively correlated with kidney function at the time of diagnostic biopsy, explained variance in gene expression (**Fig. 3h**) primarily in immune cells (14.02%), endothelial cells (EC; 11.38%), fibroblasts (FIB; 11.11%), and vascular smooth muscle/mesangial cells (vSMC/MC; 9.46%).

Pathway analysis^32^ showed a significant increase in JAK-STAT activity as Factor 5 decreased, particularly in endothelial cells (padj = 4.55e-08), together with general activation of mitogen-activated protein kinase (MAPK) pathways and nuclear factor kappa B (NF-kB) activity (**Fig. 3i**). Overrepresentation analysis of gene loadings revealed that, in PT cells, genes involved in oxidative phosphorylation (padj = 8.97e-24) and, across all cell type views, xenobiotic metabolism (padj = 8.92e-03) were more highly expressed in high-Factor 5 participants and progressively diminished in low-Factor 5 participants (**Fig. 3j**). In fibroblasts, genes related to fibroblast activation and extracellular matrix remodeling (belonging to the Hallmark Epithelial-Mesenchymal Transition gene set, padj = 1.37e-22) were upregulated as Factor 5 score decreased, suggesting a shift toward a profibrotic state.

In immune cells, genes associated with allograft rejection were downregulated in high-Factor 5 samples (padj = 2.68e-15), possibly reflecting immunosuppressive effects of the more common steroid treatment in younger participants with preserved kidney function prior to kidney biopsy, who cluster within the LowF3HighF5 quadrant (**Fig. 3f**). Although Factor 5 score is associated with steroid treatment (**Fig. 3d**), this association does not persist after stratification for pediatric participants (**SFig. S**2**b)**. Together, these results suggest that Factor 5 primarily captures transcriptional programs associated with kidney cellular metabolism which are more active in pediatric and high-eGFR participants and diminish as kidney function declines. Further, this loss of metabolic activity occurs in coordination with the establishment of a profibrotic state.

Factor 3, negatively correlated with kidney function, described transcriptional changes in the tubular compartment and explained variance particularly within the DCT (7.08%) and PT (8.74%) (**Fig. 3h**). As Factor 3 score increased, the injury markers WAP four-disulfide core domain protein 2 (WFDC2), prominin 1 (PROM1), thymosin beta 4 x-linked (TMSB4X), thymosin beta 10 (TMSB10), vimentin (VIM), vascular cell adhesion molecule 1 (VCAM1), and hepatitis A virus cellular receptor 1 (HAVCR1) were upregulated in PT (**Fig. 3k**), consistent with a transition towards a stress and repair phenotype. In DCT, increased Factor 3 scores were associated with upregulation of many of these injury markers (WFDC2, PROM1, TMSB4X) and downregulation of differentiation markers uromodulin (UMOD) and transient receptor potential cation channel, subfamily M, member 6 (TRPM6), supporting a loss of epithelial identity.

Beyond intrinsic tubular injury signatures, Factor 3 described fibroblast-mediated collagen signaling in tubular cell types as indicated by ligand receptor interaction scores (right panel, **Fig. 3k**). Overrepresentation analysis revealed widespread upregulation of epithelial-mesenchymal transition (EMT)-associated genes across all cell types with an increasing Factor 3 score (**Fig. 3m**; padj = 6.26e-05). Pathway enrichment analysis further indicated that inflammatory pathways—JAK-STAT, NF-κB, and tumor necrosis factor alpha (TNF-α)—were significantly upregulated across all cell types in high-Factor 3 participants (**Fig. 3i**; padj = 1.06e-02, padj = 1.03e-03, padj = 5.34e-04 respectively), and TGF-β was significantly upregulated in the majority of cell types (9 of 13 padj < 0.05). The Factor 3 score was also positively correlated with immune cell abundance (R = 0.63, padj = 1.04e-12) and fibroblast abundance (R = 0.42, padj = 2.04e-05) (**Fig. 3l**). These findings suggested that Factor 3 reflected an injury-driven, multicellular transcriptional program characterized by tubular stress, immune activation, and progressive fibrosis, all associated with worsening kidney function.

### Mapping cell-specific effects - APOL1 expression is associated with kidney function in FSGS

Unsupervised analysis of the OCEAN snRNAseq data established JAK-STAT activation as a key disease pathway associated with kidney function decline. Since JAK-STAT activation is known to induce APOL1 mRNA expression^33,34^, which was not originally included as a feature in MCFA model training, but is one of the strongest genetic risk factors for developing nephrotic syndrome, we examined how APOL1 expression progressed from LowF3High5 through to HighF3LowF5. APOL1 expression and distribution across cell types in OCEAN identified the highest level of APOL1 expression in the nuclei of podocytes, followed by endothelial cells, and with lower levels detected in nuclei of other cell types (**Fig. 4a**). Within podocytes, APOL1 expression was highest in nuclei from participants with FSGS ( **Fig. 4b**). In pseudobulk podocyte profiles, where sufficient sequenced nuclei were available to make a reliable gene quantitation (≥20, STable S2), APOL1 was significantly higher in patients with FSGS in comparison to profiles from all other patients as well as living donor and transplant biopsy profiles (p<0.0001, one tailed t-test). In pairwise disease-comparison analyses, APOL1 was also significantly elevated in FSGS (**SFig3a**). Among the pseudobulk podocyte profiles from participants with FSGS, APOL1 increased significantly between LowF3HighF5 (higher kidney function; lower JAK-STAT activation) and HighF3LowF5 (lower kidney function; higher JAK-STAT activation) groups (**Fig. 4c**; p = 0.041, one tailed t-test).

**Figure 4.**
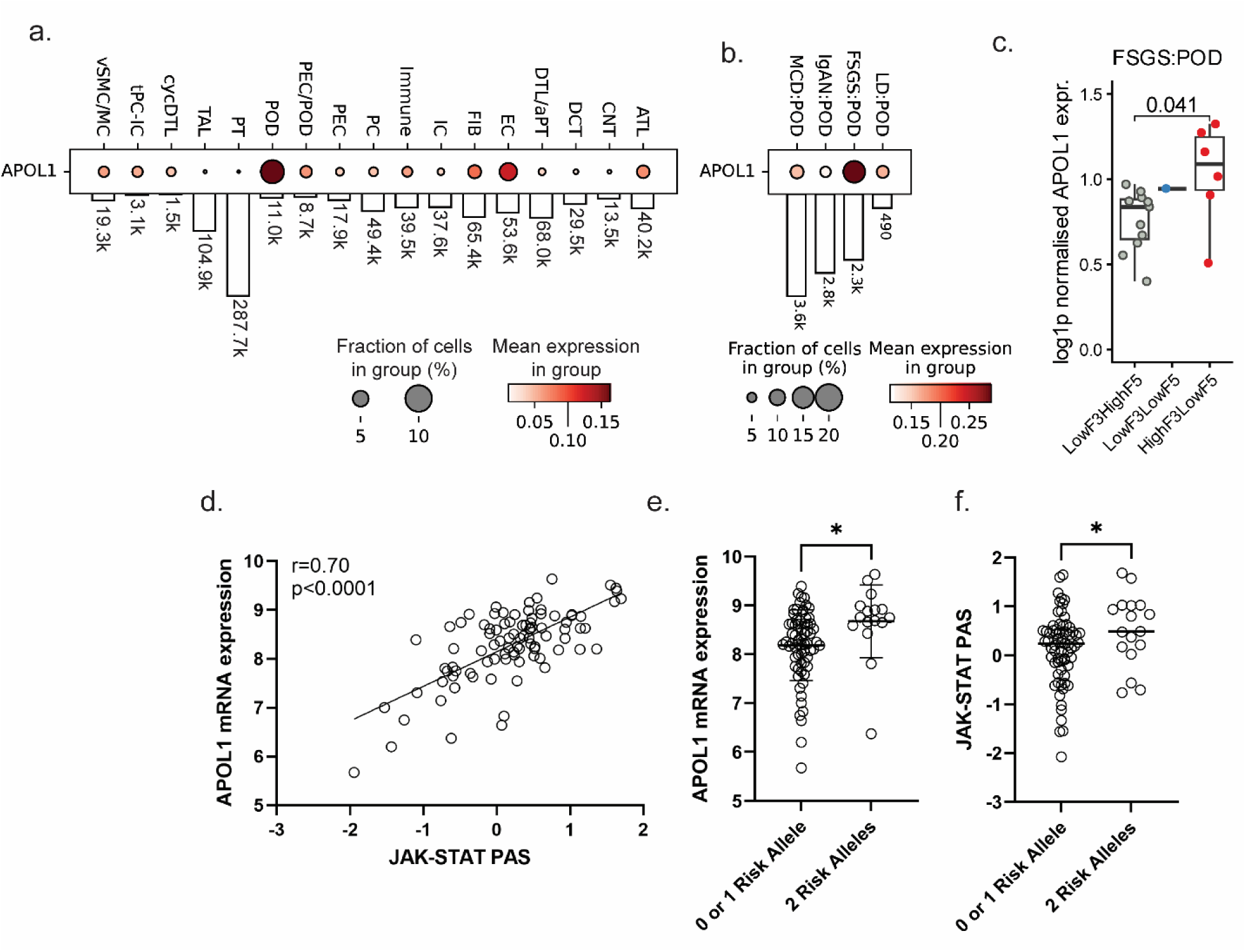
APOL1 expression is highest in podocytes from participants with FSGS and with 2 APOL1 high risk alleles. **a.** APOL1 expression, per nephron cell type, from the 10XsnRNAseq data of all participants included in OCEAN. **b.** APOL1 expression from the snRNAseq data of podocytes from living donors and participants with FSGS, IgAN, and MCD. **c.** APOL1 expression from the pseudobulked snRNAseq data of podocytes from participants with FSGS, across the LowF3HighF5, LowF3LowF5 and HighF3LowF5 groups. One tailed t-test. **d.** Pearson correlation of bulk glomerular RNAseq data from NEPTUNE participants whereby a JAK-STAT pathway activity score (PAS)^9,22^ was positively correlated with APOL1 mRNA expression. **e.** Glomerular bulk RNAseq data of APOL1 mRNA expression and **f.** JAK-STAT PAS as stratified from NEPTUNE participants with 0-1 APOL1 high risk alleles vs 2 APOL1 high risk alleles, as defined in Table 2. * = p < 0.05.

**Table 2:** Summarized Characteristics of High-Risk Cases and High-Risk Controls.

| Characteristics | High-Risk Cases<br>(N=27) | High-Risk Controls<br>(N=30) |
| --- | --- | --- |
| Mean age in years (SD, range) | 41.1 (13.4, 21-62) | 61.2 (9.8, 35-74) |
| Percent Female | 44.4% | 70% |
| Self-identified Black race | 100% | 100% |
| APOL1 Genotype (%) |  |  |
| *G1G1* | (N=16; 59.3%) | (N=13; 43.3%) |
| *G1G2* | (N= 9; 33.3%) | N=14; 46.7%) |
| *G2G2* | (N= 2; 7.4%) | (N=3; 10%) |
| Mean UPCR g/g (SD, range) | 3.1 (4.8, 0.2-18) | 0.09 (0.04, 0.05-0.2) |
| Mean eGFR (SD, range) | 35.8 (32.0, 3-103) | 90.6 (16.6, 64-124) |

Abbreviations: SD – standard deviation; APOL1- apolipoproteinL1; eGFR – estimated glomerular filtration rate; UPCR – urine protein to creatinine ratio

### Mapping cell-specific effects - APOL1 expression is correlated with JAK-STAT activation in participants with FSGS, and is elevated in those with two APOL1 high risk alleles

As we previously observed JAK-STAT activation in patients with FSGS and IgAN^11,35^, and observed the association between Factor 3/Factor 5 scores with both JAK-STAT activity and APOL1 expression in FSGS podocytes, we anticipated an association between JAK-STAT activity and APOL1 expression in bulk RNAseq profiles of glomeruli from participants with FSGS (participant clinical and demographic information included in **STable S3**) that could be assessed using the previously published transcriptional fingerprint JAK-STAT pathway activity score (PAS)^11,35^. As APOL1 was originally a component of the JAK-STAT PAS, we recomputed the JAK-STAT PAS without APOL1 to enable correlation with APOL1 mRNA expression. The computed JAK-STAT PAS with APOL1 was virtually identical to JAK-STAT PAS computed without APOL1 (r=0.998, p<0.0001, **SFig3b**), thus JAK-STAT PAS (without APOL1) performed as a reasonable surrogate for our originally described JAK-STAT PAS^11,35^, and was used for all subsequent analyses.

As expected, JAK-STAT PAS was strongly correlated with APOL1 expression in glomerular RNAseq profiles from participants with FSGS (r = 0.70, p < 0.0001, **Fig. 4d**). Participants with 2 APOL1 risk alleles had higher APOL1 mRNA expression and JAK-STAT PAS and therefore we investigated the potential association between risk allele status and concordant activation of JAK-STAT and APOL1. At the nuclei level and pseudobulk podocyte level, there was no allele specific difference detectable, possibly a consequence of the small sample size captured with a sufficient number of podocytes in OCEAN. However, in bulk RNAseq profiles available in NEPTUNE^10^, participants with 2 risk alleles (n = 16) had the highest APOL1 mRNA expression (p < 0.05) compared to those with 1 (n = 13) or 0 (n = 54) risk alleles (**Fig. 4e**), similar to previous observations^12^. Participants with 0 and 1 risk alleles had similar APOL1 mRNA expression levels, in pairwise comparisons, participants with 2 risk alleles (n = 16) had significantly higher expression than participants with 0 risk alleles (n = 54) (p < 0.05) (**SFig3c**). Similarly, the JAK-STAT PAS was highest in participants with 2 risk alleles (p < 0.05) (**Fig. 4f**), with significant difference between 0 and 2 risk alleles (p < 0.01) and an intermediate score for a single risk allele (**SFig2d**). Taken together, high APOL1 expression in podocytes in FSGS, is coupled with JAK-STAT activity suggestive of coordinated regulation of JAK-STAT activity and APOL1 regulation in FSGS.

Based on observing the gene expression profile supporting the relationship between JAK-STAT, APOL1, and APOL1 risk allele status, we proceeded to directly test the relationship between APOL1 and JAK-STAT directly in a podocyte model system.

### Functional validation in cell models: Clinical and demographic characteristics of APOL1 study participants

To test the mechanistic link between APOL1 and JAK-STAT signaling and investigate the role of this mechanism in the incomplete penetrance of APOL1-mediated kidney disease (AMKD), we established an independent case-control cohort for an iPSC-derived podocyte (iPOD) model of APOL1-mediated FSGS. We enrolled Black adults with biopsy-proven FSGS who also carried the APOL1 HR genotype, HR Cases (N = 27) and healthy Black volunteers (N=30) without diabetes, proteinuria (UPCR > 0.2 g/g)^36,37^ or loss in kidney function (eGFR < 60 ml/min, (**Fig. 5a**, **Table 2, STable S4**) for the HR control group. To increase the likelihood that an HR control is a true control, we preferentially enrolled participants 50 years or older, since APOL1-mediated FSGS becomes evident by age 50 in most cases^20,38^ (**Table 2; STable S4**). By contrast, HR cases were younger, with a mean age of 41 years, consistent with early age of onset of APOL1-mediated FSGS. HR cases also had significant proteinuria and varying degrees of eGFR decline. The high frequency of G1 allele and low frequency of G2 allele among both HR cases and HR controls were aligned with prior publications^20,39^ (**Table 2; STable S4**). There were slightly more men among the HR cases while more women had been enrolled as HR controls (**Table 2**).

**Figure 5.**
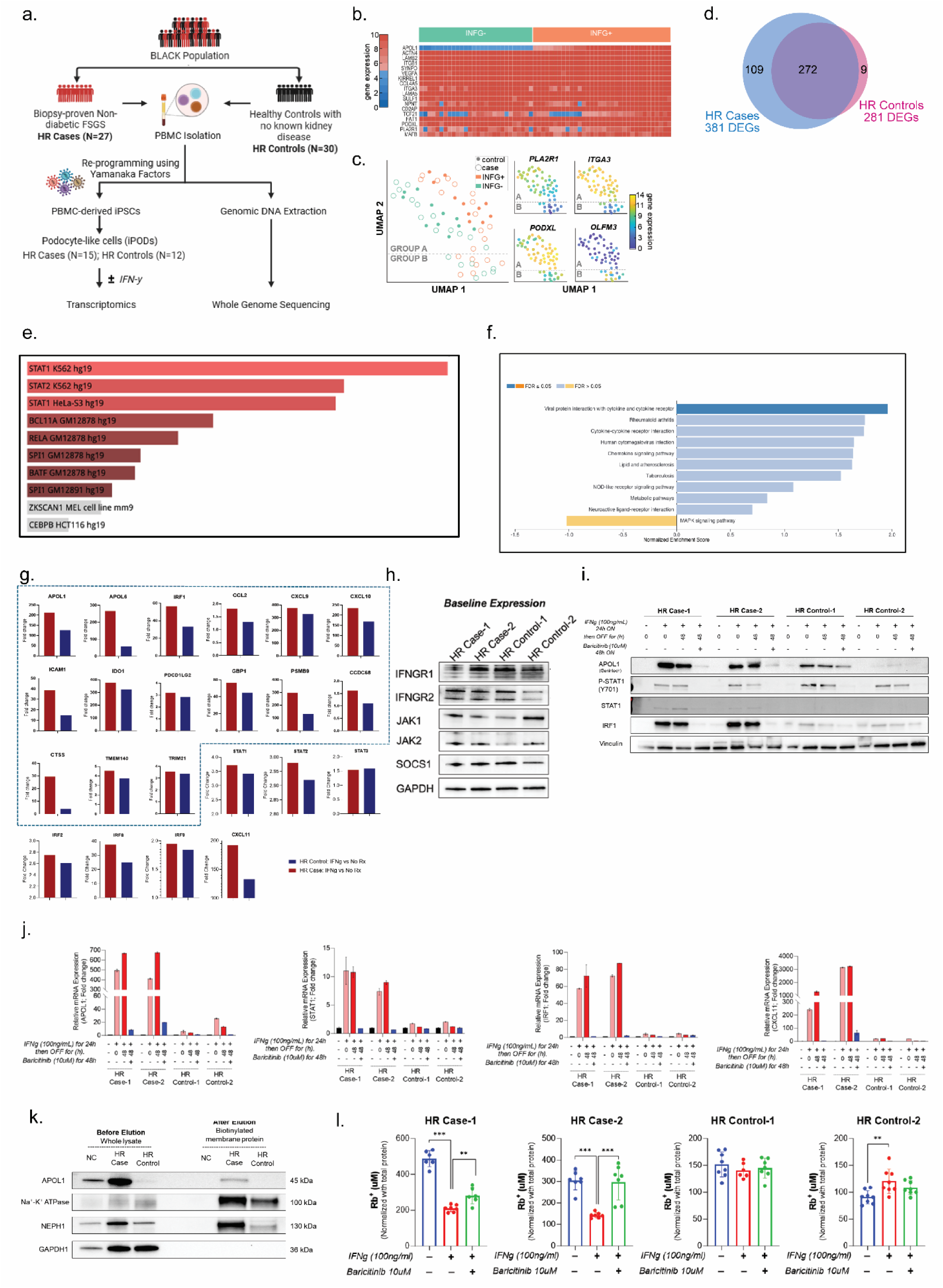
Multimodal analysis reveals increased JAK-STAT-APOL1 signaling in iPSC-derived podocytes (iPODS) of HR cases compared to HR controls. **a.** Overview of study design showing PBMC collection from participants with high-risk APOL1 genotype and biopsy-proven FSGS (HR cases, N=27) and healthy controls with high-risk APOL1 genotype but without kidney disease (HR controls, N=30). iPODs were generated from the first 15 HR cases and 12 HR controls, followed by whole genome RNA sequencing. Whole genome sequencing was performed on all 57 samples. **b.** Heat map of key podocyte-marker genes shows expression across representative iPODS of HR cases and HR controls; expression of APOL1 is induced by interferon gamma (IFNγ) treatment. **c.** UMAP partitions iPODS into two groups—successfully differentiated iPODS, group A, have good expression of podocyte markers, while poorly differentiated group B express low levels of podocyte markers. **d.** IFNγ induces 381 and 281 DEGs (padj < 0.05; fold change ≥ 2) in HR cases and HR controls, respectively, with 272 overlapping DEGs. **e.** Bar chart of top enriched terms in KEGG_2021_Human gene set library. The top 10 enriched terms for the input gene set are displayed based on the -log10(p-value). The top five enriched terms have the most significant overlap with the input query gene set, with p-values of 0.00005, 0.0002, 0.0007, 0.0001, 0.002, and 0.003, respectively. **f.** Bar chart of top enriched terms from the ENCODE_TF_ChIP-seq_2015 gene set library. The top 10 enriched terms for the input gene set are displayed based on the -log10(p-value). The top three enriched terms have the most significant overlap with the input query gene set, with p-values of 8.5^-7^, 9.6^-6^, 1.3^-5^, respectively. **g,** IFNγ induces higher mRNA fold-expression of APOL1, components of JAK-STAT signaling, and well-established STAT1-regulated genes (in dotted box) which reflects STAT1 activity in iPODs of HR cases compared to HR controls. **h.** In the first two HR cases and HR controls, the basal protein levels of JAK-STAT components are similar. **i.** However, IFNγ induces higher protein levels of APOL1, STAT1 and IRF1 in iPODs of HR cases, which are reversed by JAK inhibitor, baricitinib. **j.** Consistent with protein expression, IFNγ induces higher mRNA levels of APOL1, STAT1, IRF1 and CXCL11 in HR cases relative to HR controls (n= 2 biological replicates per group). The effects were reversed by baricitinib. **k.** membrane protein biotinylation assay shows that IFNγ-treated iPODs of HR case1 exhibit increased APOL1 localization to the plasma membrane compared to HR control1. **l.** Consistent with this finding, IFNγ-induced APOL1-mediated rubidium (Rb+) efflux in iPODs of HR cases, which were partially or completely reversed by baricitinib. All data are represented as mean ± SD. *P ≤ 0.05; ***P ≤ 0.001; ****P ≤ 0.0001, ordinary 1-wayANOVA with Tukey’s multiple-comparison test.

### Functional validation in cell models - iPSC-derived Podocytes (iPODs) Express Podocyte Markers and Respond to IFN-***γ***

To discover the dysregulated signaling pathways that distinguish podocytes of HR cases from those of HR controls, we differentiated participant derived induced pluripotent stem cells (iPSCs) from both groups into podocyte-like cells (iPODs)^40^. iPODs retained the complete genetic endowment of each participant, enabling capture of participant-specific molecular signatures with high fidelity. iPODs of both HR cases and HR controls also expressed key podocyte markers, supporting their use as podocytes surrogates (**Fig. 5b**). Similar to human primary podocytes, stimulation of iPODs with interferon gamma (IFN-γ), a known inducer of APOL1 expression via JAK1/2 kinase activation, caused upregulation of APOL1 expression across all samples (**Fig. 5b**), with clustering of untreated and IFN-γ-treated iPODs into two distinct groups (**Fig. 5c**). Quality control assessment separated iPODs into Group A and Group B based on marker expression profiles, with particular attention to podocalyxin like (PODXL), phospholipase A2 receptor 1 (PLA2R1), integrin subunit alpha 3 (ITGA3). Group B iPODs had low expression of podocyte markers (PLA2R1, PODXL and ITGA3) and high expression of olfactomedin 3 (OLFM3), suggesting Group B iPODs were poorly differentiated (**Fig. 5c**).

Therefore, the Group B iPODs were excluded from further analysis. Taken together, these results demonstrated that the iPODs expressed characteristic podocyte markers, responded to IFN-γ stimulation, and exhibited variability in APOL1 expression influenced by genetic risk factors.

### Functional characterization of JAK-STAT-APOL1 interaction in cell models - JAK-STAT Signaling is Potentiated in HR Cases and Leads to Higher APOL1 Expression

To investigate the differential response to IFN-γ stimulation, gene expression profiles were analyzed in HR cases and HR controls under both baseline and stimulated conditions.

Comparative analysis showed that under IFN-γ stimulation, there were 381 differentially expressed genes (DEGs, padj <0.05 and fold change ≥ 2) in HR cases of which 362 genes were upregulated and 19 genes downregulated. In HR controls, 281 DEGs with 277 genes upregulated and 4 genes downregulated were observed (**Fig. 5d**). Of these, 272 DEGs were common to both HR Cases and HR Controls. Importantly, 109 DEGs were unique to HR cases, of which 94 genes were upregulated and 15 downregulated. Analysis of the 94 upregulated genes using ENCODE TF ChIP-Seq data revealed STAT1 (padj < 0.0006) and STAT2 (padj < 0.003), key components of JAK-STAT pathway (**Fig. 5e**), as the top enriched transcription factors. Pathway enrichment analysis of the 94 upregulated genes using using KEGG (**Fig. 5f**) demonstrated significant enrichment of immune and inflammatory signaling pathways, including upregulation of pro-inflammatory cytokines, cytokine receptors and chemokines. These include CXCL2 (MIP-2a) and TNFRSF1B (TNFR2) reported to be associated with faster progression of kidney disease among individuals with high-risk APOL1 genotypes^41^. Consistent with these findings, individual analysis of key validated STAT1-regulated genes (dashed box, Fig. 5g)^11,42^ and JAK-STAT pathway-associated genes—STAT1, STAT2, IRF1, IRF2, and IRF8—revealed higher expression levels in HR cases relative to HR controls (**Fig. 5g**). Notably, a trend towards higher APOL1 expression was evident in HR cases compared to HR controls further supporting JAK-STAT driving activation in APOL1 HR cases.

Baseline analysis revealed that the levels of JAK-STAT components, including total STAT1, were similar between HR cases and HR controls, as shown in **Fig. 5h**. Comparative analysis of the whole genome of HR cases and HR controls did not identify variants in JAK-STAT genes or other genes that reached genome-wide significance (**SFig. S4**). However, upon stimulation with IFN-γ, HR cases exhibited higher STAT1 and IRF1 mRNA and protein (**Fig. 5i and j, SFig.S5**). Notably, the phosphorylation of STAT1, a critical activation step, did not appear to be markedly increased in HR cases versus controls, pointing to alternative mechanism(s) for the enhanced transcriptional activation of target genes in HR cases. Further investigation demonstrated elevated expression of IFN-γ-responsive genes, including CXCL11, reinforcing the heightened sensitivity of HR cases to IFN-γ signaling (**Fig. 5j**). Notably, treatment with the JAK inhibitor baricitinib effectively reversed the IFN-γ -induced expression of STAT1, IRF1, and APOL1 in HR cases (**Fig. 5i and j**), thereby confirming that the observed effects are triggered downstream of JAK1/2 kinase activation.

### Functional validation in cell models - APOL1 Protein Localization and Monovalent Cation Transport in iPODs

We and others previously reported that APOL1 G1 or G2-mediated transport of K and Na across the plasma membrane is the proximal driver of APOL1-mediated cytotoxicity including podocytopathy^43–45^. Here, we determined that the APOL1 protein localized prominently on the plasma membrane of iPODs derived from HR cases, correlating with the elevated APOL1 expression levels (**Fig. 5k**). To study ion transport dynamics, rubidium (Rb) was employed as a tracer for potassium (K) due to its physicochemical similarity and suitability for transport assays^46,47^. We ascertained that iPODs of HR cases displayed significantly higher levels of Rb efflux compared to HR control samples, indicating enhanced ion transport activity associated with increased APOL1 expression (**Fig. 5l**). Furthermore, treatment with baricitinib, partially reversed the elevated Rb efflux in HR case iPODs (**Fig. 5l**). This suggested that the APOL1-mediated ion transport is modulated, at least in part, by JAK-dependent signaling pathways, presenting a potential therapeutic target for addressing APOL1-associated cellular dysfunction in podocytes.

### Functional validation in cell models - APOL1-knockout in HR iPODs attenuated JAK-STAT signaling and abolished IFNγ-induced Rb^+^ efflux

To explore the hypothesis that APOL1 risk variants activate JAK-STAT signaling, we used CRISPR/Cas9 to knock out APOL1 in iPODs generated from a participant with G1G2 APOL1 genotype with diagnosis of FSGS (**Fig. 6a**). We confirmed lack of IFN-γ-induced APOL1 protein expression by immunofluorescence and immunoblot (**Fig. 6b and c**). IFN-γ induced robust expression of APOL1 and other interferon regulated genes that are components of the JAK-STAT pathway as confirmed by qPCR (**Fig. 6d**) and RNAseq (**Fig. 6e**). Consistent with lack of APOL1 protein, the APOL1-knockout iPODs failed to efflux rubidium compared to APOL1-competent, isogenic control iPODs (**Fig. 6f**).

**Figure 6.**
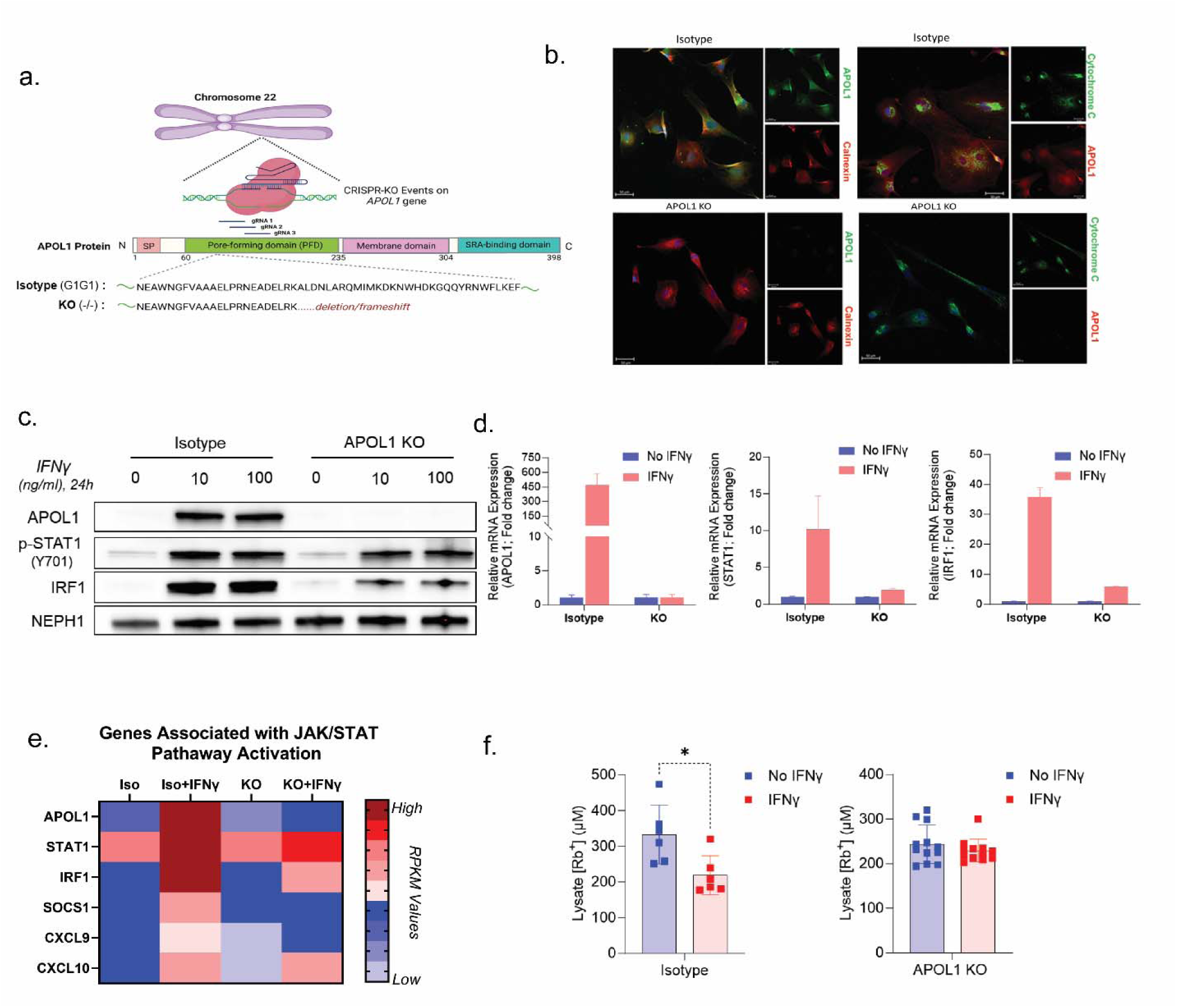
CRISPR/Cas9-mediated knockout of APOL1 attenuates JAK-STAT signaling and rubidium efflux in iPOD of a HR case. **a.** Overview of CRISPR/Cas9-mediated knockout of APOL1 in iPSC derived from HR Case1. **b.** Compared with isogenic control, iPODs derived from APOL1 knockout (KO) iPSC lack detectable APOL1 by immunofluorescence staining following treatment with IFNγ. **c**. IFNγ-induced STAT phosphorylation and IRF1 expression is reduced in APOL1 KO iPOD. **d.** Consistently, IFNγ-induced expression of STAT1 and IRF1 mRNA is also attenuated in APOL1 KO iPOD. **e.** Heat map shows reduction in IFNγ-induced, JAK-STAT regulated genes (mRNA). **f.** Unlike isogenic control iPODs in which IFNγ-induced APOL1 causes Rb+ efflux, there are no Rb+ efflux APOL1-KO IPODs (n= 10 biological replicates per group). All data are represented as mean ± SD. *P ≤ 0.05, 2-tailed t-test.

### Validation of potentiated JAK-STAT pathway in HR cases by comparing with NEPTUNE cohort

Lastly, we evaluated the 94 genes up-regulated in IFN-γ-induced cases, that were enriched for STAT1 and STAT2 target genes (**Fig. 5e**) for concordant expression changes in the glomerular biopsy transcriptome of patients with FSGS; 87 genes were detected in the glomerular transcriptome of patients with FSGS and 26 were significantly differentially regulated in FSGS compared to LD (p-adj<0.05). Of the 26 differentially regulated genes, 19 (73%) were concordantly regulated (ie. genes that were selectively up-regulated in IFN-γ iPODs from APOL1 HR cases were also up-regulated in FSGS compared to LD), while only 7 genes were discordantly regulated (ie. genes that were selectively up-regulated in IFN-γ iPODs from APOL1 HR cases were down-regulated in FSGS compared to LD). These data support translation of the IFN-γ stimulation in iPSC podocytes *ex vivo* to the APOL1-JAK-STAT interaction observed *in vivo*.

Taken together, these results strongly support the model where cytokine activation of JAK-STAT induces APOL1, which further acts in a feed forward manner to potentiate JAK-STAT signaling as evidenced by the strong correlation of JAK-STAT PAS and APOL1, in addition to higher JAK-STAT PAS in patients with two APOL1 risk alleles. Ablation of APOL1 activity through targeted knockdown of APOL1, suppresses JAK-STAT dependent signaling consistent with this model (**Fig. 7**).

**Figure 7.**
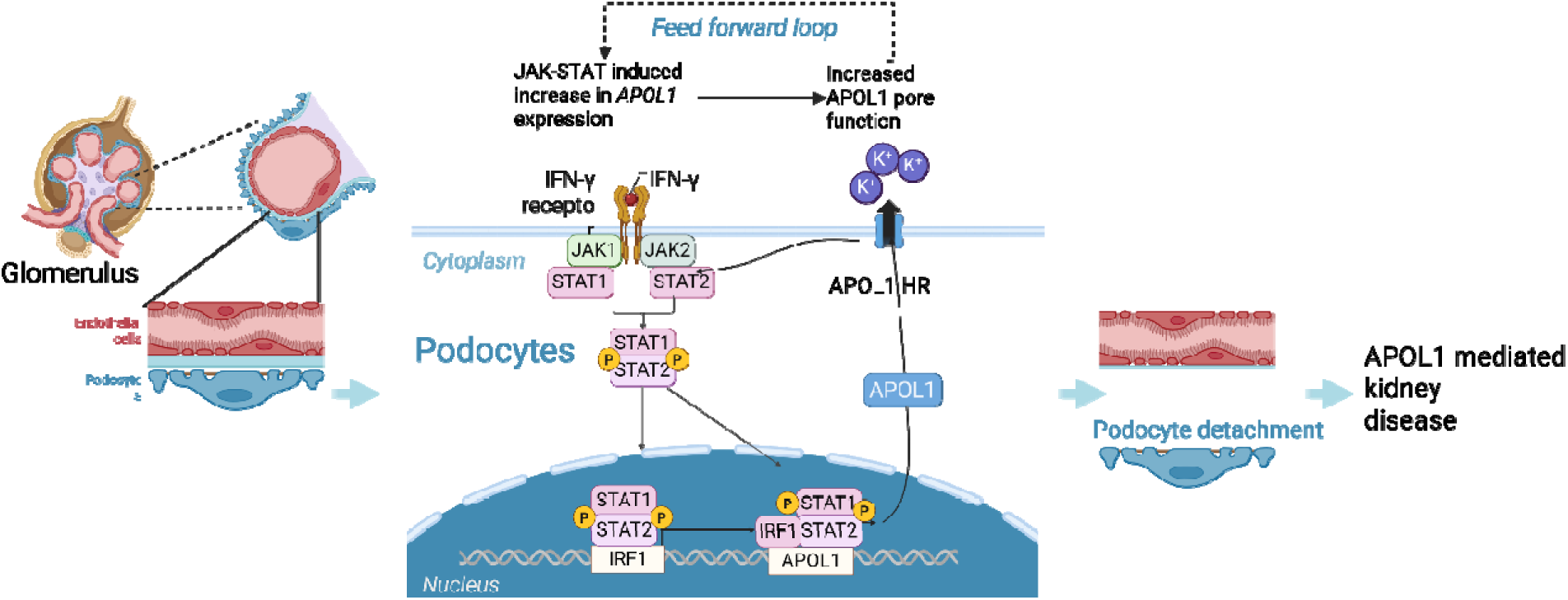
Summary schematic. Proposed model of JAK-STAT induced APOL1 expression feed forward loop on progression towards APOL1 mediated kidney disease. “JAK-STAT-APOL1 Feed Forward Model” created in BioRender. by Subramanian, L. and is licensed under CC BY 4.0.

## Discussion

Here, we report the generation of OCEAN, a large-scale single cell and single-nucleus transcriptomic rare glomerular diseases atlas for the community to investigate cell-level effects to address a variety of different questions, including cellular changes with disease trajectory and across disease entities leading to potential cell selective markers for evaluating kidney health.

This dataset and the subsequent use case with APOL1 demonstrate the power of molecular disease definitions in rare glomerular disease to bring novel therapies into the clinic. We illustrated the utility of coupling large scale single nucleus-level profiling with existing transcriptomic data and analysis suites to glean insight into underlying mechanisms in rare kidney diseases and association with clinical and genetic phenotypes. While an exhaustive analysis of cell types and their role in rare kidney disease is beyond the scope of this manuscript, OCEAN can be effectively used as a resource for the kidney research community to enhance mechanistic understanding of disease.

To define the key underlying determinants of glomerular disease on a cell resolved level we deployed a multicellular factor analysis (MCFA) framework^30^. MCFA allowed unbiased mapping of the core determinants of observed variance in OCEAN in terms of multi-cellular factors within different cell-types distinguishing biology from any technical variation. These factors were interpreted based on the underlying molecular mechanisms such as pathways and cell-cell communication. By correlating these factors to patient-level phenotypic and clinical characteristics, we identified general and etiology-specific drivers of disease progression. Factors associated with kidney damage, showed enrichment of molecular programs known to be active in progressive loss of kidney function, more broadly, but now resolved at the cellular level in rare glomerular diseases. Demonstrating the activation of these disease progression programs to be in a disease stage but not disease-type specific manner is a critical insight for the glomerular disease community, as it provides the rational to include primary glomerular diseases into umbrella trials cutting across conventional disease categories, rather targeting shared mechanism of disease (NCT05003986)^28,48^.

Localization of the expression of APOL1 predominantly in podocytes and endothelial cells led to the identification of the JAK-STAT pathway, characterized by activation of its downstream molecular signature, and its strong association with APOL1 expression in the glomeruli, particularly in patients with FSGS, prompted further cell-type specific mechanistic evaluations. iPSC-derived podocytes from people with APOL1-associated FSGS showed increased JAK-STAT activation in response to IFN-γ stimulation, suggesting that potentiated JAK-STAT signaling may be a modifier of AMKD. Furthermore, CRISPR/Cas9-mediated knockout of APOL1 in iPODs, attenuated JAK-STAT activation, suggesting an APOL1 pore-dependent, feed forward mechanism.

Chronic JAK-STAT activation in the context of APOL1 could explain the elevated APOL1 expression observed in participants with HR APOL1 compared to those without such that once activated, it becomes a disease driving feed-forward loop. The limited self-renewing capacity of nephron progenitor cells into podocytes, compared to endothelial or tubular epithelial cells^49,50^, may explain the unique vulnerability of podocytes to this pathological feed-forward loop.

What is clear from these findings and from epidemiological studies^39,51^ is that APOL1 HR genotype is not sufficient for AMKD. Presumably other genetic, epigenetic, or environmental factors leading to activated JAK-STAT pathways might also be necessary. It remains to be seen if other cytokine or inflammatory pathways (e.g. IL-1β or TNF activation) could also serve as initiators of disease activity leading to a chronic activation state. It also remains to be seen if elevated APOL1 expression itself may be a contributor to AMKD in the absence of APOL1 HR, but in the presence of JAK-STAT activation.

While single-cell and single-nucleus data from participants are observational and correlative in nature, the introduction of iPODs, and prior studies in kidney organoids,^10,33^ demonstrate a clear causal relationship between IFN-γ stimulation to JAK-STAT activation to induction of APOL1 mRNA expression. Although iPODs may not fully recapitulate mature podocyte behavior *in vivo*, the recapitulation of human disease biology strongly suggests that, at least in relation to the JAK-STAT-APOL1 regulatory loop, the model functions as intended.

Further, the marked differences between iPODs from APOL1 HR cases and controls suggests potential applications in patient-centric AMKD risk stratification or drug-response assessment in precision nephrology, not unlike patient-derived xenografts in oncology^52^.

To further strengthen this feed-forward model, it will be important to test whether APOL1 risk variants alone are sufficient to drive JAK/STAT activation in the absence of cytokine stimulation. Experimental strategies such as CRISPRa-mediated induction or overexpression of G1/G2 versus G0 alleles could directly address this question. Importantly, published *in vivo* work^19,53^ provides supportive evidence: doxycycline-induced expression of APOL1 risk variants in mouse podocytes, independent of exogenous interferon, was sufficient to enhance JAK/STAT signaling and promote glomerular injury. These findings align with our proposed model and highlight APOL1 risk variants as active amplifiers of JAK/STAT signaling rather than passive downstream effectors. The data presented herein supports the mechanistic use of JAK-STAT^54^ and APOL1 inhibitors^55^ in APOL1 high risk patients. The ongoing JUSTICE trial (NCT05237388) of baricitinib in AMKD is likely to reveal whether inhibition of JAK-STAT signaling is an effective therapeutic strategy.

In summary, we have created a comprehensive kidney tissue transcriptional atlas focused on glomerular diseases that is freely available to the glomerular disease research community to explore (Neptune-ocean.cxg.miktmc.org). This atlas provides a foundation for treatment target and disease biomarker discovery in patients with MCD, FSGS, and IgAN. Specifically, we demonstrated the utility of the atlas in identifying disease-associated transcriptional networks in FSGS by providing a human cell–type resolved mechanistic link between APOL1 risk variants and JAK/STAT-driven podocyte injury in FSGS. This mechanistic link is now being targeted in the ongoing JUSTICE trial, part of the precision medicine trial platform of NEPTUNE Match^28^, which closes the loop from molecular profiling to targeted therapeutic intervention for glomerular diseases.

## Methods

### Participant cohorts - NEPTUNE and HKTTA

NEPTUNE (NCT01209000) is a prospective observational study that enrolls children and adults with proteinuria at the time of their first clinically indicated kidney biopsy from 21 participating sites^1^. Human Kidney Transplant Transcriptomic Atlas (HKTTA) is a prospective cohort of kidney transplant recipients, who were willing to provide an additional biopsy core for research purposes at the time of clinical biopsy pre- and post-transplantation. OCEAN included snRNAseq and scRNAseq data from kidney biopsies of 120 NEPTUNE participants and 50 HKTTA participants. An additional 92 NEPTUNE participants with FSGS with previously published bulk-RNAseq profiles^2^ were included in the analysis. All clinical and research activities reported here are consistent with the principles of the Declaration of Istanbul.

### Duke University and Partners HealthCare study participants for functional validation

In accordance with protocols approved by Partners HealthCare System and Duke University Health System IRB, self-identified Black or African American adults (age 18 years or older) with biopsy-proven focal segmental glomerulosclerosis (FSGS, N = 51) were consented and enrolled from nephrology clinics in Boston, Massachusetts and Durham, North Carolina between 2019 and 2023. Among these participants, 27 were found to have a APOL1 high-risk (HR) genotype. We labeled these participants high-risk (HR) cases. We consented healthy, self-identified Black or African American adults (and 50 years or older) with no known kidney disease (N=218). We enrolled the 30 healthy volunteers (N=30) found to have high-risk APOL1 genotype as high-risk (HR) controls. We excluded volunteers with an eGFR less than 60ml/min/1.73 m^2^, a UPCR > 0.2, a low-risk APOL1 genotype, or found to have diabetes mellitus. The minimal enrollment age of 50 years for controls was informed by prior reports that indicate that APOL1-associated, clinically significant kidney disease typically manifests by age 50^3–5^. All, except one HR control participant, met this age cutoff. Individuals in the HR control group were also excluded if they had proteinuria (urine protein-to-creatinine ratio [UPCR] > 0.2 g/g) or an estimated glomerular filtration rate (eGFR) < 60 mL/min/1.73 m², or diabetes mellitus.

### Whole genome sequencing of NEPTUNE participants

Blood samples provided by consenting NEPTUNE participants were subjected to whole genome sequencing; reads were obtained using Illumina HiSeq. Sequencing was performed to an average read depth of at least 30X. FASTQ files were then aligned to the GRCh38 draft of the human genome using bwa-mem (version 0.7.17). Joint variant calling and filtering was performed using GATK (version 4.1.8.0) following recommended best practices (https://gatk.broadinstitute.org/hc/en-us/articles/360035535932-Germline-short-variant-discovery-SNPs-Indels). The genotype calls for the following APOL1 variant were then extracted for participants having snRNAseq data: G1: rs73885319, rs60910145, G2: rs71785313, and N264K: rs73885316. Participants without any calls for the G1 or G2 variants were noted as having 0 risk alleles.

### Isolation of single nuclei for snRNAseq and single cells for scRNAseq

Nuclei from RNAlater-preserved biopsies of 120 NEPTUNE and 11 HKTTA participants, were processed as described previously^2^. Cells from Cryostor-preserved biopsies of 50 HKTTA participants, were isolated according to a published protocol^6^). All HKTTA biopsy samples, including the 11 HKTTA samples that were processed for snRNAseq (dx.doi.org/10.17504/protocols.io.86khzcw), were processed for scRNAseq (dx.doi.org/10.17504/protocols.io.7dthi6n). Some participants in HKTTA contributed kidney biopsy samples at different time points within a year.

### 10x sample processing

10x snRNAseq and scRNAseq were performed following standard protocols, using the 10x Chromium Single-Cell 3′ Reagent Kit v3 kit. Sample demultiplexing, barcode processing, and gene expression quantifications were conducted using the 10x Cell Ranger pipeline (v.3-7.1) using the GRCh38 (hg38) reference genome. For 10x snRNAseq, introns were included in Cell Ranger analyses.

### Quality control of 10XsnRNAseq and scRNAseq data

For 10x snRNAseq, nuclei and cognate barcodes that passed 10x Cell Ranger filters were used for downstream analysis in a Seurat environment (Seurat v.4.3.0.1) ^7–9^. SoupX (v.1.0.0-1.6.0) ^10^ was conducted on all samples at default parameters to remove ambient RNA contamination in samples. Nuclei containing greater than or equal to 10% of all transcripts originating from the mitochondrial genome were removed. Nuclei that had an nFeatures count <500 or >5000 were also removed to minimize severely damaged nuclei or multiplets. DoubletFinder (v.2.0.0-2.0.3) ^11^, was used at default parameters. While the 10x scRNAseq processing used SoupX and DoubletFinder as described above, cells that contained greater than or equal to 50% of all transcripts originating from the mitochondrial genome were removed. 10x scRNAseq nFeatures criteria were identical to snRNAseq processing during integration.

### Parse sample processing

Parse Biosciences snRNAseq was performed using standard protocols using the Evercode WT v2 (Mega) kit on a 48-well or 96-well plate. Nuclei that passed the Parse quality control pipeline were read into a Seurat environment. Nuclei that contained an nFeatures count less than 200 or greater than 5000 were removed. Nuclei containing only protein-coding mitochondrial transcripts greater than 10% of all transcripts were also removed.

### Integration and clustering analysis

All snRNAseq and scRNAseq datasets were integrated into one Seurat object via RPCA-based integration. Integration anchors were selected from datasets that are conditional exemplars of the entire dataset. Here, snRNAseq from FSGS, MCD, and IgAN backgrounds were used in conjunction with sn and scRNAseq in LD backgrounds (**supplementary methods**). The IntegrateData function in Seurat required k.weight=75, since several scRNAseq samples contained low cell numbers. The resulting integrated object was scaled and subjected to principal components analysis to determine the number of clusters. Each cluster was subjected to the FindAllMarkers function in Seurat, with resulting cluster markers compared, manually, to known markers for kidney cell types to infer cluster labels^6,12^. Clinical metadata, including APOL1 risk allele calls, were appended to the Seurat object using the package tidyseurat (v.0.8.0). UMAPs and imaging of differentially expressed genes was conducted either with the Seurat package or with the VIP plugin (v.3.0.0) for cellxgene (v.1.1.1) ^13^.

### Pseudobulking OCEAN snRNAseq, Bulk-RNAseq data and correlations of APOL1 and JAK-STAT activity

Glomerular cell types, namely podocytes, transitioning parietal epithelium, parietal epithelium, vascular smooth muscle/mesangium, glomerular endothelium mixed with the peritubular endothelium, and afferent/efferent arterioles, were pooled from select FSGS patients and a pseudobulked dataset was constructed using the AggregateExpression function in Seurat.

Correlations were conducted as previously published^14^. JAK-STAT transcriptional activity was computed by aggregating Z-transformed gene expression profiles of JAK-STAT target genes into an average Z-score. Removal of APOL1 from the JAK-STAT gene list was conducted when a direct comparison to APOL1 was examined.

### Post-processing of OCEAN data for unsupervised analysis of multicellular programs

OCEAN data were processed for multicellular factor analysis (MCFA) ^15^ in Python using scanpy^16^. Cluster annotations were mapped to their major cell types (**STable S1**). The dataset was subsetted to single-nucleus RNAsequencing data. Cell and gene filtering were performed using scanpy.pp.filter_cells (min_genes=200, default parameters otherwise) and scanpy.pp.filter_genes (min_cells=3, default parameters otherwise).

### Creation of multi-view pseudobulk expression object

The filtered AnnData object was converted to a MuData object containing aggregated pseudobulk expression profiles for each cell type using functionality of the decoupler package^17^. To ensure high-quality pseudobulk profiles, a minimum of 50 cells per cell type in one sample was required. Single-nucleus gene expression was aggregated into sample-level pseudobulk profiles using decoupler.get_pseudobulk (mode=’sum’, min_cells=50, min_counts=100, min_smpls=3). The pseudobulk expression profiles were further filtered at the gene level using decoupler.filter_by_expr (min_count=10, min_total_count=15, large_n=5, min_prop=0.05).

Sample filtering ensured that each sample was represented in at least two cell-type views. After gene and sample filtering, pseudobulk profiles were normalized using scanpy.pp.normalize_total (target_sum=10,000, default parameters otherwise). A variance-stabilizing transformation was applied using scanpy.pp.log1p with default parameters.

### Cell-cell communication inference

Putative ligand-receptor interaction scores were calculated using LIANA+^18^. Putative ligand-receptor interactions were identified based on co-expression using liana.mt.rank_aggregate.by_sample (expr_prop=0.1, use_raw=False, n_perms=100, return_all_lrs=False, default parameters otherwise). The resulting ligand-receptor scores were converted into a multi-view object using liana.multi.lrs_to_views (score_key=’magnitude_rank’, lr_prop=0.1, lrs_per_sample=20, lrs_per_view=20, samples_per_view=10, min_variance=0, lr_fill=0, default parameters otherwise).

### Feature selection for MulticellularFactor Analysis (MCFA)

Samples in OCEAN were collected over four sub-projects, which were found to be a source of variance within the snRNAseq data. To mitigate this, we applied batch-aware highly variable gene selection as a soft batch-correction method, focusing on variation within batches rather than inter-batch variance. Highly variable genes were identified for each cell type from the pseudobulk expression profiles using <u>sc.pp</u>.highly_variable_genes, with the sub-project (‘Project1’) as the batch_key, and default parameters otherwise. In ligand-receptor views, features were retained when at least one component (ligand or receptor) was identified as a batch-aware highly variable gene in the corresponding sender or receiver cell type.

### Variance decomposition using Multicellular Factor Analysis (MCFA)

Following feature selection, the cell type pseudobulk expression and ligand-receptor multi-view objects were concatenated into a single MuData object for input to Multicellular Factor Analysis, which was performed using fucntions of the package for MultiOmics Factor Analysis (MOFA)^19^. MOFA was run using muon.tl.mofa (use_obs=’union’, convergence_mode=’medium’, seed=1337), with 20 Factors. As feature selection had already been applied, use_var was set to None. Where not otherwise specified, default parameters were used. Factor scores and variable loadings (i.e., genes and ligand-receptor pairs) were extracted for downstream analysis using liana.utils.get_factor_scores and liana.utils.get_variable_loadings, respectively. Total R^2^ values across all samples were computed using calculate_variance_explained from the mofax package (per_factor=True, default parameters otherwise). Per-sample R^2^ values were calculated using the same function with sample as group_label (‘OCEAN_MS_Exp_ID’).

### CLR transformation

To prepare cell type compositional data for statistical testing, a centered log-ratio (CLR) transformation was applied to cell type proportions per sample using clr and multiplicative_replacement from the skbio.stats.composition module^20^.

### Enrichment analysis

Pathway activity inference on latent factor gene expression loadings were conducted using decoupler.run_ulm. Pathway activities were inferred using the PROGENy ^21^ prior knowledge on pathways’ target genes, retrieved via decoupler.get_progeny (organism=’human’, top=500, default parameters otherwise). Overrepresentation analysis (ORA) on the top and bottom 200 loadings for Factor 3 and Factor 5 using Hallmark gene sets^22^ was performed with decoupler.get_ora_df, with gene sets obtained from decoupler.get_resource(’MSigDB’) and filtered for the Hallmark collection.

### Data visualization for unsupervised patient variability analysis

All unsupervised patient variability visualizations for Figure 3 were generated in R using ggplot2^23^, ggpubr^24^, patchwork^25^ and ComplexHeatmap^26^.

### Low pass whole genome sequencing (LP-WGS) of Duke University and Partners HealthCare study participants

Low-pass whole-genome sequencing (LP-WGS) was performed in two batches by Genewiz, Azenta Life Sciences, on DNA samples from 57 African American individuals carrying APOL1 high-risk (HR) genotypes. Sequencing reads were aligned to the GRCh38 human reference genome, and variant imputation was performed by Genewiz. Variant call files (VCFs) were merged, and variants failing quality thresholds based on Genome Analysis Toolkit (GATK) Best Practices were excluded. Subsequent data processing and quality control (QC) steps were conducted using PLINK^27^. Variants with missingness >15% and those with minor allele frequency (MAF) <0.05 were excluded. To further assess sample quality and relatedness, we generated a linkage disequilibrium-pruned dataset comprising biallelic variants with MAF ≥0.30. Identity-by-descent (IBD) analysis was conducted to detect duplicate or highly related samples. Given the admixed ancestry of the cohort (predominantly African with some European ancestry), we implemented an ancestry-aware genome-wide association study (GWAS) using Tractor^28^.

Local ancestry inference (LAI) was performed with RFMix^29^ and ancestry-specific karyograms were constructed to visualize African and European ancestry tracts across the genome. Tractor was then used to perform GWAS stratified by local ancestry, producing separate association results for African and European segments. For comparison, a conventional GWAS was also performed using PLINK. In both models, sequencing batch was included as a covariate.

Manhattan plots were generated in R for visualization of GWAS results. To explore the functional relevance of top-associated variants, significant SNPs were queried against the NephQTL database^30^, which contains expression quantitative trait loci (eQTLs) derived from glomerular and tubulointerstitial tissues in the NEPTUNE cohort.

### Reprogramming of PBMCs to Induced pluripotent stem cells (iPSCs)

Peripheral blood was collected from 15 APOL1 high-risk (HR) cases and 12 HR controls. Peripheral blood mononuclear cells (PBMCs) were isolated using SepMate-50 tubes (Cat# 85460) and Lymphoprep (Cat# 07801) following manufacturer’s protocol. PBMCs were reprogrammed into iPSCs using the CytoTune™ 2.0 Sendai Reprogramming Kit (Thermo Fisher Scientific, Cat# A16517), following the manufacturer’s protocol and established guidelines. On day 0, 5 × 10 PBMCs were seeded in 1 mL of PBMC Expansion Medium and transduced with Sendai virus vectors encoding the reprogramming factors OCT3/4, SOX2, KLF4, and c-MYC (designated as KOS, hc-MYC, and hKLF4) at multiplicities of infection (MOI) of either 5:5:3 or 10:10:10. Following viral transduction, cells were spinoculated by centrifugation at 2,250 rpm for 90 minutes at room temperature. After 24 hours, the culture medium was replaced with fresh PBMC Expansion Medium. On day 1, 2 × 10 irradiated mouse embryonic fibroblasts (MEFs) were plated onto Geltrex-coated plates as a feeder layer. MEF medium was composed of DMEM/F12 + GlutaMAX, supplemented with 1% embryonic stem (ES) cell-qualified fetal bovine serum, 1% MEM non-essential amino acids (NEAA), 1% penicillin-streptomycin (all from Thermo Fisher Scientific), and 1 μL/mL of 55 mM β-mercaptoethanol (Sigma-Aldrich). On day 3, transduced PBMCs were transferred onto the MEF feeder layer, where they attached and subsequently formed iPSC colonies.

### Characterization of iPSCs

Selected iPSC lines were characterized for pluripotency markers, trilineage differentiation potential, and genomic stability.

### Immunocytochemistry for pluripotency markers

Pluripotency-associated markers—SSEA4, OCT4, SOX2, and TRA-1-60—were assessed by immunocytochemistry using the PSC 4-Marker Immunocytochemistry Kit (Thermo Fisher Scientific), following the manufacturer’s protocol. Briefly, iPSCs were washed with Dulbecco’s phosphate-buffered saline (DPBS), fixed in fixative solution for 15 minutes at room temperature (RT), and permeabilized for 15 minutes. Following a 30-minute incubation in blocking solution, cells were incubated with primary antibodies for 3 hours at RT. After three washes, cells were incubated with the appropriate secondary antibody for 1 hour at RT, followed by nuclear counterstaining with NucBlue Fixed Cell Stain (Thermo Fisher Scientific).

### Trilineage differentiation via embryoid body (EB) formation and Scorecard assay

To evaluate trilineage differentiation capacity, embryoid bodies were generated by dissociating iPSC colonies into clumps using Versene (Thermo Fisher Scientific) and transferring them to ultra-low attachment flat-bottom plates (Costar, catalog number: [insert part number]). Cells were cultured in medium containing DMEM/F12 + GlutaMAX, 20% KnockOut Serum Replacement (KSR), 10% fetal bovine serum (FBS), 1% MEM non-essential amino acids, 1% penicillin-streptomycin, and 1 μL/mL 55 mM β-mercaptoethanol (Sigma-Aldrich). Revitacell™ Supplement (Thermo Fisher Scientific) was added for the first 24 hours. Media were changed every other day for 7–10 days, until EB clumps were visibly formed. Aggregates were filtered through a 30 μm filter (Miltenyi Biotec) to remove single cells. Total RNA was extracted using the RNeasy Mini Kit (Qiagen) and converted to cDNA using the SuperScript IV First-Strand Synthesis System (Thermo Fisher Scientific). Expression of lineage-specific genes was assessed using the TaqMan hPSC Scorecard Panel (384-well format; Thermo Fisher Scientific) following the manufacturer’s instructions.

### Karyotype analysis

Genomic stability of iPSC lines was assessed using the KaryoStat™ assay (Thermo Fisher Scientific), a microarray-based platform for detecting chromosomal abnormalities at high resolution.

### CRISPR–Cas9–mediated knockout of APOL1 in human iPSCs

CRISPR–Cas9–mediated knockout of APOL1 in human iPSCs was performed using the Synthego Gene KO Kit v2 (Synthego), following the manufacturer’s protocol. Three chemically modified synthetic sgRNAs targeting APOL1 were used: sgRNA #1: 5′-TAAAGCTCTGGACAACCTTG-3′; sgRNA #2: 5′-AAGGCCAGCAGTACAGAAAC-3′; sgRNA #3: 5′-AGCTTGAGGATAACATAAGA-3′. The sgRNAs were pooled in equimolar ratios and complexed with recombinant Cas9 protein (2:1 molar ratio of sgRNA:Cas9) to form ribonucleoprotein (RNP) complexes. The RNPs were incubated at room temperature for 10–15 minutes to allow complex formation. iPSCs were harvested at ∼70–80% confluency and resuspended in Neon Resuspension Buffer R. For electroporation, 2 × 10 cells were mixed with the pre-formed RNPs and transfected using the Neon Transfection System (Invitrogen) with the following parameters optimized for human iPSCs: 1,100 V, 30 ms pulse width, and 1 pulse.

Following electroporation, cells were immediately transferred into vitronectin-coated plates containing pre-warmed Stem-Flex medium (Life Technologies; A3349401) supplemented with 10 µM Y-27632 (ROCK inhibitor) and incubated at 37°C with 5% CO. 72 hours post-electroporation cells were re-plated in multiple 96-well plates in such a way that each well of 96-wells has one single cell and allows them to grow for several days. Multiple clones were selected and confirmed the APOL1 knock-out using Sanger sequencing and western blotting. CRISPR editing efficiency and indel spectra were analyzed using the ICE (Inference of CRISPR Edits) analysis tool (Synthego). Finally, possible CRISPR OFF-target effects were eliminated using whole-genome sequencing of the selected clones and differentiated in kidney podocytes (iPODs) using the methods described below.

### Differentiation of iPSCs into iPODs

Differentiation of human iPSCs into mature kidney podocytes was performed using a directed, multi-stage protocol adapted from Musah et al. ^31,32^. iPSCs were first induced toward the intermediate mesoderm lineage by treatment with CHIR99021 (a GSK-3β inhibitor) in basal media, promoting WNT signaling activation. After approximately 4 days, cells were transitioned to nephron progenitor specification media containing Activin A and BMP7 to support renal lineage commitment. Following nephron progenitor induction, cells were exposed to a podocyte maturation medium containing a combination of VEGF, retinoic acid, and a reduced concentration of CHIR99021 to promote differentiation into mature podocytes. Over the course of 14–21 days, differentiated cells acquired typical podocyte morphology, and expressed key podocyte markers such as synaptopodin (SYNPO), nephrin (NPHS1), and WT1, confirmed via immunofluorescence staining and qRT-PCR.

### RNA isolation and quantitative real-time PCR (qPCR)

Total RNA was extracted from cultured cells using the RNeasy Mini Kit (Qiagen, Cat# 74104) according to the manufacturer’s instructions. Briefly, cells were lysed in RLT buffer containing β-mercaptoethanol, followed by homogenization through pipetting. An equal volume of ethanol was added to the lysate, and the mixture was applied to a spin column for RNA binding. After sequential washes, RNA was eluted in RNase-free water and quantified using a NanoDrop spectrophotometer (Thermo Fisher Scientific). RNA quality was confirmed by A260/A280 ratios between 1.8 and 2.1. For cDNA synthesis, 1 µg of total RNA was reverse transcribed using the High-Capacity cDNA Reverse Transcription Kit with RNase Inhibitor (Applied Biosystems, Cat# 4374966), following the manufacturer’s thermal cycling protocol: 25°C for 10 min, 37°C for 120 min, and 85°C for 5 min. Synthesized cDNA was stored at −20°C until further analysis. Quantitative PCR (qPCR) was conducted using TaqMan™ Fast Advanced Master Mix (Applied Biosystems, Cat# 4444557) and TaqMan™ Gene Expression Assays (FAM-labeled) specific to the genes of interest and housekeeping genes (e.g., GAPDH or ACTB). Each 20 µL reaction contained 10 µL master mix, 1 µL 20× gene expression assay, 2 µL cDNA (∼100 ng RNA equivalent), and 7 µL nuclease-free water. Reactions were performed in optical 96-well plates on a QuantStudio™ 6 Real-Time PCR System (Applied Biosystems) using the following thermal cycling conditions: 50°C for 2 min, 95°C for 2 min, followed by 40 cycles of 95°C for 1 sec and 60°C for 20 sec. All reactions were run in technical triplicates. Gene expression was analyzed using the ΔCt method, normalizing Ct values to the housekeeping gene. Fold changes in gene expression were calculated using the 2^−ΔCt^ or 2^−ΔΔCt^ method where appropriate.

### Western blotting

Western blot analysis was performed as previously described^33^. Following experimental treatment, cells were washed with PBS and lysed in 1× Cell Lysis Buffer (Cell Signaling Technology, #9803). Lysates were sonicated and centrifuged at 10,000 rpm for 10 minutes at 4°C to collect clarified protein supernatants. Protein concentrations were quantified using a BCA Protein Assay Kit (Pierce, 23225). Equal amounts of protein (20–30 μg) were denatured in 1× Laemmli sample buffer containing β-mercaptoethanol at 100 °C for 5 minutes, separated via SDS-PAGE on precast gels (Bio-Rad), and transferred onto PVDF membranes using the Trans-Blot Turbo Transfer System (Bio-Rad). Membranes were blocked in 5% (w/v) nonfat dry milk (LabScientific, M0842) in 1× TBST (Boston BioProducts, IBB-181) and incubated overnight at 4°C with primary antibodies. The following primary antibodies were used: APOL1 (rabbit anti-human, Genentech, 3.1C1 & 3.7D6, 1:5000, 0.05 μg/mL; mouse anti-human, Genentech, 4.17A5, 4 μg/mL for IF), sodium–potassium ATPase (Abcam, ab76020; IF 1:500, WB 1:100,000), GAPDH (mouse anti-human, Santa Cruz, sc-47724, 1:200), vinculin (mouse anti-human, Sigma-Aldrich, V9131, 1:200), Stat1 (D1K9Y) (Rabbit anti-human, CST #14994T), Phospho Stat1 (Tyr701) (58D6) (Rabbit anti-human, CST #9167S), IRF-1 (D5E4) (Rabbit anti-human, CST# 8478T), and NEPH1 (Santa Cruz Biotechnology, sc-373787). All antibodies were used at manufacturer-recommended dilutions. After washing, membranes were incubated for 1 hour at room temperature with HRP-conjugated secondary antibodies (anti-mouse IgG, CST #7076; anti-rabbit IgG, CST #7074). Signal was developed using SuperSignal West Dura Extended Duration Substrate (Thermo Fisher Scientific, #34075) and imaged with the Bio-Rad ChemiDoc MP Imaging System. Band intensities were quantified by densitometry using AlphaView SA software (ProteinSimple).

### Immunofluorescence staining

Cells were washed with phosphate-buffered saline (PBS) and fixed in 4% paraformaldehyde (PFA) in PBS for 15 minutes at room temperature. Following fixation, cells were washed three times with PBS and incubated in blocking buffer (1% bovine serum albumin in PBS) for 1 hour. Primary antibodies were applied overnight at 4°C. After washing, appropriate fluorescently labeled secondary antibodies were applied for 1 hour at room temperature. Nuclei were counterstained and mounted using ProLong™ Glass Antifade Mountant with NucBlue™ Stain (Thermo Fisher Scientific, Cat# P3698). Fluorescent images were acquired using either an ECHO Revolve microscope or a Zeiss LSM 780 upright confocal microscope.

### Cell surface protein biotinylation assay

Cell surface protein labeling and isolation were performed using the Pierce™ Cell Surface Biotinylation and Isolation Kit (Thermo Fisher Scientific, #A44390) according to the manufacturer’s instructions and described previously^3^. Briefly, cells were seeded on 6-well plates and grown to ∼95% confluency. Following IFN-γ (10ng/ml) treatment, cells were washed with PBS and incubated with EZ-Link™ Sulfo-NHS-SS-Biotin for 10 minutes at room temperature to selectively label surface-exposed primary amines. Excess reagent was quenched with ice-cold 1× TBS (provided in the kit). Cells were subsequently lysed in ice-cold lysis buffer (provided in the kit) for 30 minutes. Total lysates were collected, and 5% of the input was reserved for comparison in immunoblotting. Biotinylated proteins were isolated using NeutrAvidin™ resin, and elution of enriched surface proteins was carried out following the kit protocol. Western blotting was then performed using both total protein input (5%) and eluted biotin-labeled cell surface fractions. APOL1 expression was assessed in both fractions, with Na /K ATPase used as a plasma membrane loading control and GAPDH used as a control for total input lysates.

### Measurement of Rb flux in iPODs

Rb flux, a surrogate for potassium transport, was measured in human iPSC-derived podocytes (iPODs) using X-ray fluorescence (XRF)–based quantification. A total of 25,000 iPODs were seeded onto laminin-521–coated 96-well plates and treated with interferon-γ (IFN-γ, 10 ng/mL) in the presence or absence of baricitinib (10 μM) for 45 hours. Cells were subsequently incubated with 5 mM rubidium chloride (RbCl), a potassium analog, for 3 hours to allow intracellular uptake. After incubation, cells were washed three times with modified 1× Earle’s Balanced Salt Solution (EBSS) lacking Ca² and K to remove extracellular Rb, then air-dried. Intracellular Rb content was quantified using XRpro® X-ray fluorescence technology, as previously described^3^. All data are represented as mean ± SD. *P ≤ 0.05; ***P ≤ 0.001; ****P ≤ 0.0001. 2-tailed t-test and ordinary 1-wayANOVA with Tukey’s multiple-comparison test were used for statistical analysis.

## Data Availability

All biopsy samples processed directly for inclusion in this study will be uploaded to the Gene Expression Omnibus (GEO) under accession number and publicly available upon publication. The constructed data object is currently hosted online at Neptune-ocean.cxg.miktmc.org, and will be deposited in the CellXGene server upon publication81. Biopsy samples previously uploaded to GEO that were included in this composite dataset and manuscript can be found at GSE213030 (snRNAseq) and GSE254957 (Bulk RNAseq). Whole genome sequencing calls of APOL1 will be made available.

https://neptune-ocean.cxg.miktmc.org/

## Acknowledgements

We would like to thank Lalita Subramanian for help with writing, editing and graphics for this paper. Library prep and next-generation sequencing was carried out in the Advanced Genomics Core at the University of Michigan. The Nephrotic Syndrome Study Network (NEPTUNE) is part of the Rare Diseases Clinical Research Network (RDCRN), which is funded by the National Institutes of Health (NIH) and led by the National Center for Advancing Translational Sciences (NCATS) through its Division of Rare Diseases Research Innovation (DRDRI). NEPTUNE is funded under grant number U54DK083912 as a collaboration between NCATS and the National Institute of Diabetes and Digestive and Kidney Diseases (NIDDK). Additional funding and/or programmatic support is provided by the University of Michigan, NephCure Kidney International, Alport Syndrome Foundation, and the Halpin Foundation. RDCRN consortia are supported by the RDCRN Data Management and Coordinating Center (DMCC), funded by NCATS and the National Institute of Neurological Disorders and Stroke (NINDS) under U2CTR002818. This study was supported, in part, by the George M. O’Brien Michigan Kidney Translational Resource Center (MKTC), funded by NIH/NIDDK grant U54DK137314 to MK. This study was also supported by research grants from Maze therapeutics, the NEPTUNE Public private partnership involving Travere, Boehringer Ingelheim, Sanofi and Dimerix. The human stem cell-derived podocyte model of APOL1 nephropathy project was funded by NIH Director New Innovators Award to OAO, grant number 1DP2DK124891 and by 5R01MD016401. Additional funding was provided by Massachusetts General Hospital, Duke University, and the German Science Foundation (DFG) funding through the Clinical Research Unit 5011 InteraKD (Project ID: 445703531 to JSR). For computational analysis, the authors acknowledge support by the state of Baden-Württemberg through bwHPC and the German Research Foundation (DFG) through grant INST 35/1597-1 FUGG, as well as the data storage service SDS@hd supported by the Ministry of Science, Research and the Arts Baden-Württemberg (MWK) and the German Research Foundation (DFG) through grant INST 35/1503-1 FUGG.

## Author contributions

SE, PJM, CB, SD, OAO, JS-R, MK conceived and planned the study; SE, PJM, EAO, SD, SM, KS, DF, JH, FA, FAA, BG, LHM, ASN performed the experiments or acquired data; SE, PJM, EAO, FAA, VN, KZL, MA, CB, RF, AEA-K, MEG, SD, OAO, ML analyzed the data; SE, PJM, CB, SD, OAO, VN, MTM, DL, MGS, JS-R interpreted the findings; SE, PJM, CB, SD, OAO, VN, JH, JS-R, MK edited and wrote the manuscript. All authors reviewed and approved the manuscript.

## Competing interest declaration

MK reports grants and contracts through the University of Michigan outside of this work from Chan Zuckerberg Initiative, AstraZeneca, NovoNordisk, Eli Lilly, Boehringer-Ingelheim, European Union Innovative Medicine Initiative, Certa Therapeutics, RenalytixAI, Regeneron, Novo Nordisk, Sanofi, Dimerix, Travere and Vera Therapeutics. He has received consulting fees through the University of Michigan from NovoNordisk, Alexion, Novartis, Roche Diagnostics and Vera Therapeutics. MK and VN have a patent PCT/EP2014/073413 “Biomarkers and methods for progression prediction for chronic kidney disease” licensed. MK has served on the NIH-NCATS council, is the committee chair for the American Society of Nephrology Program and is on the board of Nephcure Kidney International. SE receives research funding through the University of Michigan from AstraZeneca PLC, Eli Lilly and Company, NovoNordisk, and Travere Therapeutics, Gilead Sciences Inc, Moderna Inc, and IONIS Pharmaceuticals Inc. OAO received research funding through Duke University from Icagen, and an investigational study drug from Eli Lilly. He received consulting fees from Maze and Podium Bio. JS-R reports in the last 3 years funding from GSK and Pfizer and fees/honoraria from Travere Therapeutics, Stadapharm, Astex, Owkin, Pfizer, Grunenthal, Tempus and Moderna. All other authors have no conflicts to report.

## Materials and correspondence

Correspondence and material requests should be addressed to Sean Eddy

## List of Supplementary Materials

- List of contributors for the NEPTUNE study
- Supplementary Tables

- STable S1. Cluster annotation for the major cell types in the MOFA view
- STable S2 – Number of podocytes sequenced per patient with FSGS
- STable S3 - Characteristics of NEPTUNE participants with FSGS and bulk glomerular transcriptomic data
- STable S4 – Demographic and clinical characteristics of case-control cohort for iPSC-derived podocyte (iPOD) model of APOL1 mediated FSGS
- Supplementary Figures

- SFigS1 - UMAPs by disease category/group and Seurat specific cluster output
- SFigS2 – Clinical information from NEPTUNE participants stratified by Factor 3-Factor 5 status.
- SFigS3 - JAK-STAT-APOL1 expression in podocytes of participants with FSGS.
- SFigS4 - Lack of genetic modifiers in HR cases and HR controls
- SFigS5 - Interferon regulated downstream effects in iPODS from HR cases and HR controls

## Notes

### Clinical Trial

NCT01209000

### Author Declarations

The NEPTUNE and HKTtA studies and associated activities are approved by the ethic committee/IRB at the University of Michigan. The APOL1 validation study and protocols are approved by Partners HealthCare System and Duke University Health System IRB

### Summary of Updates

Additional author contributions and re-evaluation of N264K mutation status in NEPTUNE participants.

