## Supplemental Files for "A Human Glomerular Disease Atlas defines the APOL1-JAK-STAT feed forward loop in focal segmental glomerulosclerosis"

**List of Supplementary Materials**

- List of contributors for the NEPTUNE study
- Supplementary Tables
  - STable S1. Cluster annotation for the major cell types in the MOFA view
  - STable S2 – Number of podocytes sequenced per patient with FSGS
  - STable S3 - Characteristics of NEPTUNE participants with FSGS and bulk glomerular transcriptomic data
  - STable S4 – Demographic and clinical characteristics of case-control cohort for iPSC-derived podocyte (iPOD) model of APOL1 mediated FSGS
- Supplementary Figures
  - SFigS1 - UMAPs by disease category/group and Seurat specific cluster output
  - SFigS2 – Clinical information from NEPTUNE participants stratified by Factor 3-Factor 5 status.
  - SFigS3 - JAK-STAT-APOL1 expression in podocytes of participants with FSGS.
  - SFigS4 - Lack of genetic modifiers in HR cases and HR controls
  - SFigS5 - Interferon regulated downstream effects in iPODS from HR cases and HR controls

### Members of the Nephrotic Syndrome Study Network (NEPTUNE)

#### NEPTUNE Collaborating Sites

*Atrium Health Levine Children's Hospital, Charlotte, SC*: Susan Massengill[*], Layla Lo[#]
*Cleveland Clinic, Cleveland, OH*: Katherine Dell[*], John O'Toole[*], John Sedor[**], Victoria Grange[#]
*Children's Hospital, Denver, CO:* Bradley Dixon[*], Nathan Rogers[#]
*Children's Hospital, Los Angeles, CA*: Rachel Lestz[*], Natalie Esquivias[#]
*Children's Mercy Hospital, Kansas City, MO*: Tarak Srivastava[*], Kelsey Markus[#]
*Cohen Children's Hospital, New Hyde Park, NY*: Christine Sethna[*], Suzanne Vento[#]
*Columbia University, New York, NY:* Pietro Canetta[*]
*Duke University Medical Center, Durham, NC:* Opeyemi Olabisi[*], Rasheed Gbadegesin[**], Maurice Smith[#]
*Emory University, Atlanta, GA:* Laurence Greenbaum[*], Chia-shi Wang[*], Chris Fan[#]
*The Lundquist Institute, Torrance, CA:* Sharon Adler[*], Janine LaPage[#]
*John H Stroger Cook County Hospital, Chicago, IL:* Amatur Amarah[*]
*Johns Hopkins Medicine, Baltimore, MD:* Meredith Atkinson[*], Ryan Hutson[#]
*Mayo Clinic, Rochester, MN:* John Lieske, Marie Hogan, Fernando Fervenza
*Medical University of South Carolina, Charleston, SC:* David Selewski[*], Cheryl Alston[#]
*Montefiore Medical Center, Bronx, NY:* Kim Reidy[*], Michael Ross[*], Frederick Kaskel[**], Patricia Flynn[#]
*New York University Medical Center, New York, NY:* Laura Malaga-Dieguez[*], Olga Zhdanova[**], Laura Jane Pehrson[#], Melanie Miranda[#]
*The Ohio State University College of Medicine, Columbus, OH*: Salem Almaani[*], Laci Roberts[#]
*Riley Children's Hospital of Indiana University, Indianapolis, IN:* Myda Khalid[*], Veronica Servin[#]
*Stanford University, Stanford, CA:* Richard Lafayette[*], Elizabeth Chen[#]
*Temple University, Philadelphia, PA:* Iris Lee[**]
*Texas Children's Hospital at Baylor College of Medicine, Houston, TX*: Shweta Shah[*], Thinh Phan[#]
*University Health Network Toronto:* Heather Reich[*], Michelle Hladunewich[**], Paul Ling[#], Martin Romano[#]
*University of California at San Diego, San Diego, CA:* Ambarish Athavale[*], Caitlin Carter[*], Kristin Zeeb[#]
*University of California at San Francisco, San Francisco, CA*: Paul Brakeman[*], Daniel Schrader
*University of Colorado Anschutz Medical Campus, Aurora, CO*: James Dylewski[*] Nathan Rogers[#]
*University of Kansas Medical Center, Kansas City, KS*: Ellen McCarthy[*], Catherine Creed[#]
*University of Miami, Miami, FL:* Alessia Fornoni[*], Miguel Bandes[#]
*University of Michigan, Ann Arbor, MI:* Matthias Kretzler[*], Laura Mariani[*], Zubin Modi[*], Amanda Williams[#], Roxy Ni[#]

*University of Minnesota, Minneapolis, MN:* Patrick Nachman[*], Michelle Rheault[*], Ariel Langenberger[#], Brady Wallner[#]
*University of North Carolina, Chapel Hill, NC:* Vimal Derebail[*], Keisha Gibson[*], Anne Froment[#], Sharia Warren[#]
*University of Pennsylvania, Philadelphia, PA:* Lawrence Holzman[*], Kevin Meyers[**], Krishna Kallem[#], Arielle Swenson[#]
*University of Texas San Antonio, San Antonio, TX*: Samin Sharma[**]
*University of Texas Southwestern, Dallas, TX:* Elizabeth Roehm[*], Kamalanathan Sambandam[**], Elizabeth Brown[**]
*University of Washington, Seattle, WA:* Ashley Jefferson[*], Sangeeta Hingorani[**], Katherine Tuttle[**§], Linda Manahan [#], Emily Pao[#], Kelli Kuykendall[§]
*Wake Forest University Baptist Health, Winston-Salem, NC:* Jen Jar Lin[**]
*Washington University in St. Louis, St. Louis, MO*: Brian Stotter[*], Joseph Dumayas[#]

**Data Analysis and Coordinating Center:** *University of Michigan:* Matthias Kretzler[*], Brenda Gillespie[**], Laura Mariani[**], Zubin Modi[**], Eloise Salmon[**], Howard Trachtman[**], Hailey Desmond, Sean Eddy, Damian Fermin, Wenjun Ju, Maria Larkina, Chrysta Lienczewski, Rebecca Scherr, Jonathan Troost, Amanda Williams, Yan Zhai; *Cleveland Clinic:* Crystal Gadegbeku[**], John Sedor[**], *Duke University:* Laura Barisoni[**]; *Harvard University:* Matthew G Sampson[**]; *Northwestern University:* Abigail Smith[**]; *University of Pennsylvania:* Lawrence Holzman[**], Jarcy Zee[**]

**Digital Pathology Committee:** Carmen Avila-Casado *(University Health Network)*, Serena Bagnasco *(Johns Hopkins University)*, Lihong Bu *(Mayo Clinic)*, Shelley Caltharp *(Emory University)*, Clarissa Cassol *(Arkana)*, Dawit Demeke *(University of Michigan)*, Brenda Gillespie *(University of Michigan)*, Jared Hassler *(Temple University)*, Leal Herlitz *(Cleveland Clinic)*, Stephen Hewitt *(National Cancer Institute)*, Jeff Hodgin *(University of Michigan)*, Danni Holanda *(Arkana)*, Neeraja Kambham *(Stanford University)*, Kevin Lemley, Laura Mariani *(University of Michigan)*, Nidia Messias *(Washington University)*, Alexei Mikhailov *(Wake Forest)*, Vanessa Moreno *(University of North Carolina)*, Behzad Najafian *(University of Washington)*, Matthew Palmer *(University of Pennsylvania)*, Avi Rosenberg *(Johns Hopkins University)*, Virginie Royal *(University of Montreal)*, Miroslav Sekulik *(Columbia University)*, Barry Stokes *(Columbia University)*, David Thomas *(Duke University)*, Ming Wu *(University of New York)*, Michifumi Yamashita *(Cedar Sinai)*, Hong Yin *(Emory University)*, Jarcy Zee *(University of Pennsylvania)*, Yiqin Zuo *(University of Miami)*. Co-Chairs: Laura Barisoni *(Duke University)*, Cynthia Nast *(Cedar Sinai)*.

[*]Principal Investigator; [**]Co-investigator; [#]Study Coordinator; [§]Providence Medical Research Center, Spokane, WA Last Update: 19NOV2024

**STable S1. Gene expression profiles aggregated at the major cell type level**

| Cluster annotation | Major cell type - MFA view |
|---|---|
| ATL | ATL |
| CNT | CNT |
| cycDTL | DTL/aPT |
| DCT | DCT |
| DTL/aPT | DTL/aPT |
| EC | EC |
| FIB | FIB |
| IC | IC |
| Immune | Immune |
| vSMC/MC | vSMC/MC |
| PC | PC |
| DTL | DTL/aPT |
| PEC | PEC/POD |
| POD | PEC/POD |
| PT | PT |
| TAL | TAL |
| tPC-IC | PC |

**STable S2 – Podocyte counts of sequenced nuclei in OCEAN from NEPTUNE participants with FSGS**

| ID | # Sequenced Podocytes | Sequencing Technology | APOL1 risk alleles | ID | # Sequenced Podocytes | Sequencing Technology | APOL1 risk alleles |
|---|---|---|---|---|---|---|---|
| PT105 | 215 | 10x | 0 | PT59 | 37 | 10x | 0 |
| PT106 | 7 | Parse | 0 | PT61 | 17 | 10x | 0 |
| PT109 | 6 | 10x | 2 | PT61 | 16 | Parse | 0 |
| PT109 | 5 | Parse | 2 | PT67 | 12 | 10x | 2 |
| PT112 | 516 | 10x | 2 | PT67 | 6 | Parse | 2 |
| PT112 | 5 | Parse | 2 | PT69 | 20 | 10x | 2 |
| PT113 | 13 | Parse | 0 | PT70 | 2 | 10x | 2 |
| PT120 | 25 | 10x | 1 | PT72 | 8 | 10x | 1 |
| PT120 | 34 | Parse | 1 | PT74 | 21 | 10x | 2 |
| PT128 | 3 | Parse | 0 | PT82 | 1 | Parse | 2 |
| PT130 | 23 | 10x | 2 | PT86 | 107 | Parse | 2 |
| PT130 | 3 | Parse | 2 | PT97 | 44 | 10x | 2 |
| PT136 | 5 | 10x | 0 | PT97 | 3 | Parse | 2 |
| PT136 | 1 | Parse | 0 | | | | |
| PT26 | 51 | 10x | 0 | | | | |
| PT26 | 10 | Parse | 0 | | | | |
| PT30 | 58 | 10x | 2 | | | | |
| PT30 | 50 | Parse | 2 | | | | |
| PT32 | 44 | 10x | 1 | | | | |
| PT32 | 41 | Parse | 1 | | | | |
| PT35 | 29 | 10x | 1 | | | | |
| PT37 | 233 | 10x | 0 | | | | |
| PT37 | 46 | Parse | 0 | | | | |
| PT38 | 287 | 10x | Not measured | | | | |
| PT43 | 1 | 10x | 2 | | | | |
| PT43 | 1 | Parse | 2 | | | | |
| PT44 | 53 | 10x | 2 | | | | |
| PT44 | 36 | Parse | 2 | | | | |
| PT48 | 29 | 10x | 0 | | | | |
| PT49 | 55 | 10x | 2 | | | | |
| PT49 | 14 | Parse | 2 | | | | |
| PT51 | 4 | Parse | 0 | | | | |
| PT52 | 423 | 10x | 2 | | | | |
| PT55 | 5 | 10x | 0 | | | | |
| PT57 | 81 | 10x | 1 | | | | |
| PT57 | 6 | Parse | 1 | | | | |

**STable S3. Characteristics of NEPTUNE participants with FSGS and bulk glomerular transcriptomic data**

| Characteristic | FSGS |
|---|---|
| Number | 92 |
| Mean age in years ±SD (range) | 32.7±22.0 (2-77) |
| % Female | 37 |
| % Pediatric | 37 |
| **Number self-reported race*** | |
| Asian/Asian American | 4 |
| Black/African American | 36 |
| Multi-racial | 3 |
| White/Caucasian | 48 |
| Not reported | 7 |
| **Clinical factors** | |
| % APOL1 high risk | 22 |
| Mean eGFR at baseline ± SD(mL/min 1.73m2) | 73.6±31.1 |
| UPCR at baseline (median, IQR) | 2.4 (1.0, 4.8) |

*Race options provided were: Asian/Asian American; Black/African American; Multi-Racial; Native American/Alaskan Native/First Nation; Native Hawaiian/Other Pacific Islander; White/Caucasian; Unknown. Missing and unknown were combined under “not reported”.

**STable S4 – Demographic and clinical characteristics of case-control cohort for iPSC-derived podocyte (iPOD) model of APOL1 mediated FSGS**

| Case/ control ID | Age Range | Gender | Genotype | Diagnosis | UPCR[a] range at enrollment | Serum Creatinine[b] at enrollment |
|---|---|---|---|---|---|---|
| Case1 | 40-49 | F | G1G1 | Collapsing FSGS | 0.3-1.0 | 0.8 |
| Case 2 | 20-29 | F | G1G1 | FSGS | 1.0-3.0 | 11.53 |
| Case 3 | 50-59 | M | G1G2 | FSGS | 1.0-3.0 | 7.28 |
| Case 4 | 40-49 | M | G1G2 | FSGS | >3 | ESRD |
| Case 5 | 50-59 | F | G1G1 | Collapsing FSGS | 0.3-1.0 | 2.1 |
| Case 6 | 20-29 | M | G1G1 | FSGS | 1.0-3.0 | 2.86 |
| Case 7 | 20-29 | M | G1G2 | Collapsing FSGS | 1.0-3.0 | 2.6 |
| Case 8 | 20-29 | M | G1G2 | Collapsing FSGS | 0.3-1.0 | 1 |
| Case 9 | 30-39 | M | G2G2 | FSGS | 1.0-3.0 | 95 |
| Case 10 | 20-29 | F | G1G2 | Collapsing FSGS | 1.0-3.0 | 1.1 |
| Case 11 | 30-39 | M | G1G1 | FSGS | >3 | 9.9 |
| Case 12 | 20-29 | F | G1G1 | FSGS | >3 | 2.9 |
| Case 13 | 60-69 | F | G1G1 | FSGS | N/A | 12.63 |
| Case 14 | 50-59 | M | G1G2 | Collapsing FSGS | 1.0-3.0 | 3.5 |
| Case 15 | 50-59 | M | G1G1 | FSGS | 0.3-1.0 | 9.55 |
| Case 16 | 50-59 | M | $G1^{GM}G1^{G+}$ | FSGS | N/A | ESRD |
| Case 17 | 30-39 | F | G1G1 | FSGS | N/A | ESRD |
| Case 18 | 40-49 | M | G1G1 | FSGS, ESRD | 1.0-3.0 | ESRD |
| Case 19 | 40-49 | M | G1G1 | Collapsing FSGS | >3 | 1.5 |
| Case 20 | 50-59 | M | G1G1 | FSGS | N/A | 1.53 |
| Case 21 | 60-69 | F | G1G1 | Collapsing FSGS | 0.3-1.0 | 1.36 |
| Case 22 | 20-29 | F | G1G2 | FSGS | 0-0.3 | 0.8 |
| Case 23 | 50-59 | M | G1G2 | FSGS | N/A | 12.18 |
| Case 24 | 30-39 | F | G1G1 | FSGS | N/A | 8.95 |
| Case 25 | 20-29 | F | G2G2 | FSGS | N/A | 1.1 |
| Case 26 | 20-29 | M | G1G1 | FSGS | N/A | 12.08 |
| Case 27 | 50-59 | F | G1G2 | FSGS | 1.0-3.0 | 1.83 |
| Control 1 | 30-39 | M | G1G2 | Healthy control | 0-0.3 | 0.69 |
| Control 2 | 50-59 | M | G1G1 | Healthy control | 0-0.3 | 1.1 |
| Control 3 | 50-59 | F | G1G2 | Healthy control | 0-0.3 | 0.85 |
| Control 4 | 50-59 | F | G1G1 | Healthy control | 0-0.3 | 0.93 |
| Control 5 | 60-69 | F | G1G1 | Healthy control | 0-0.3 | 1 |
| Control 6 | 70-79 | M | G1G1 | Healthy control | 0-0.3 | 1.06 |

| | | | | | | |
|---|---|---|---|---|---|---|
| Control 7 | 50-59 | M | G1G1 | Healthy control | 0-0.3 | 0.91 |
| Control 8 | 60-69 | M | G1G2 | Healthy control | 0-0.3 | 0.79 |
| Control 9 | 60-69 | F | G1G1 | Healthy control | N/A | 0.64 |
| Control 10 | 50-59 | F | G1G2 | Healthy control | 0-0.3 | 0.86 |
| Control 11 | 50-59 | F | G1G2 | Healthy control | 0-0.3 | 1.11 |
| Control 12 | 60-69 | M | G1G1 | Healthy control | N/A | 1.13 |
| Control 13 | 50-59 | F | G1G2 | Healthy control | 0-0.3 | 0.79 |
| Control 14 | 70-79 | F | G1G1 | Healthy control | 0-0.3 | 0.94 |
| Control 15 | 70-79 | M | G1G2 | Healthy control | 0-0.3 | 1.02 |
| Control 16 | 70-79 | F | G2G2 | Healthy control | 0-0.3 | 0.77 |
| Control 17 | 60-69 | F | G1G2 | Healthy control | 0-0.3 | 0.61 |
| Control 18 | 60-69 | F | G1G1 | Healthy control | 0-0.3 | 1.02 |
| Control 19 | 50-59 | M | G1G1 | Healthy control | 0-0.3 | 0.94 |
| Control 20 | 50-59 | F | G1G2 | Healthy control | 0-0.3 | 0.84 |
| Control 21 | 50-59 | F | G1G2 | Healthy control | 0-0.3 | 0.8 |
| Control 22 | 70-79 | M | G1G1 | Healthy control | N/A | 0.98 |
| Control 23 | 60-69 | F | G1G2 | Healthy control | 0-0.3 | 0.73 |
| Control 24 | 50-59 | F | G2G2 | Healthy control | 0-0.3 | 0.85 |
| Control 25 | 70-79 | F | G1G2 | Healthy control | 0-0.3 | 0.82 |
| Control 26 | 70-79 | F | G1G2 | Healthy control | 0-0.3 | 0.96 |
| Control 27 | 70-79 | F | G2G2 | Healthy control | 0-0.3 | 0.53 |
| Control 28 | 50-59 | F | G1G1 | Healthy control | 0-0.3 | 0.84 |
| Control 29 | 70-79 | F | G1G1 | Healthy control | 0-0.3 | 0.95 |
| Control 30 | 50-59 | F | G1G2 | Healthy control | 0-0.3 | 0.74 |

[*]Urine protein to creatinine ratio (g/g) is reported as a range and [b]serum creatinine measured in g, both at time of enrollement

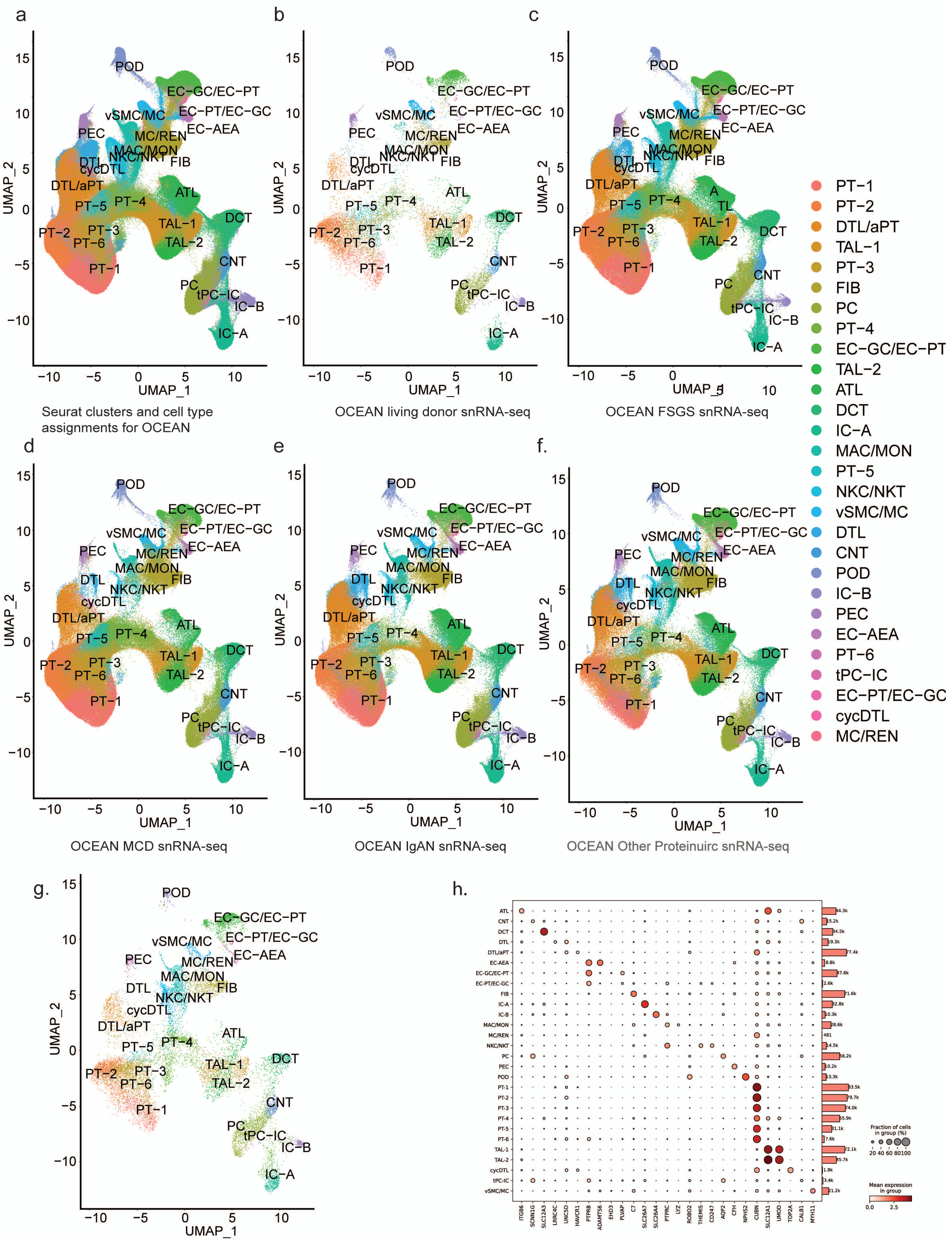


**SFig.S1. UMAPs by disease category/group and Seurat specific cluster output. a.** Twenty eight clusters were derived from the rpca-based integration of every dataset contained within OCEAN, which are characterized here. EC-GC = glomerular endothelium; EC-PT = peritubular endothelium; IC-A = type a intercalated cells; IC-B = type b intercalated cells; MAC/MON = myeloid-derived immune cells; NKC/NKT = natural killer cells/natural killer T cells; EC-AEA = afferent/efferent arterioles; MC/REN = mesangial, renin-positive, cells that may also contain juxtaglomerular cells. For instances where more than one cluster of a specific cell type was characterized, sequential iteration was noted for each additional characterization of the same cell type (e.g., PT-1, PT-2, etc.) For proximal tubules (PT) cell types, this is not additional commentary on the S1, S2, or S3 compartments. The EC-GC/EC-PT cluster contains both cell types, though likely has more glomerular endothelial cells than peritubular endothelial cells; the converse is true for the EC-PT/EC-GC cluster. **b-g.** UMAP projections depicting the nuclei and their distribution from a specific condition, as indicated in the figure caption. **h**. Cell type markers that were used in determining the cell type assignments for the Seurat-derived clusters.

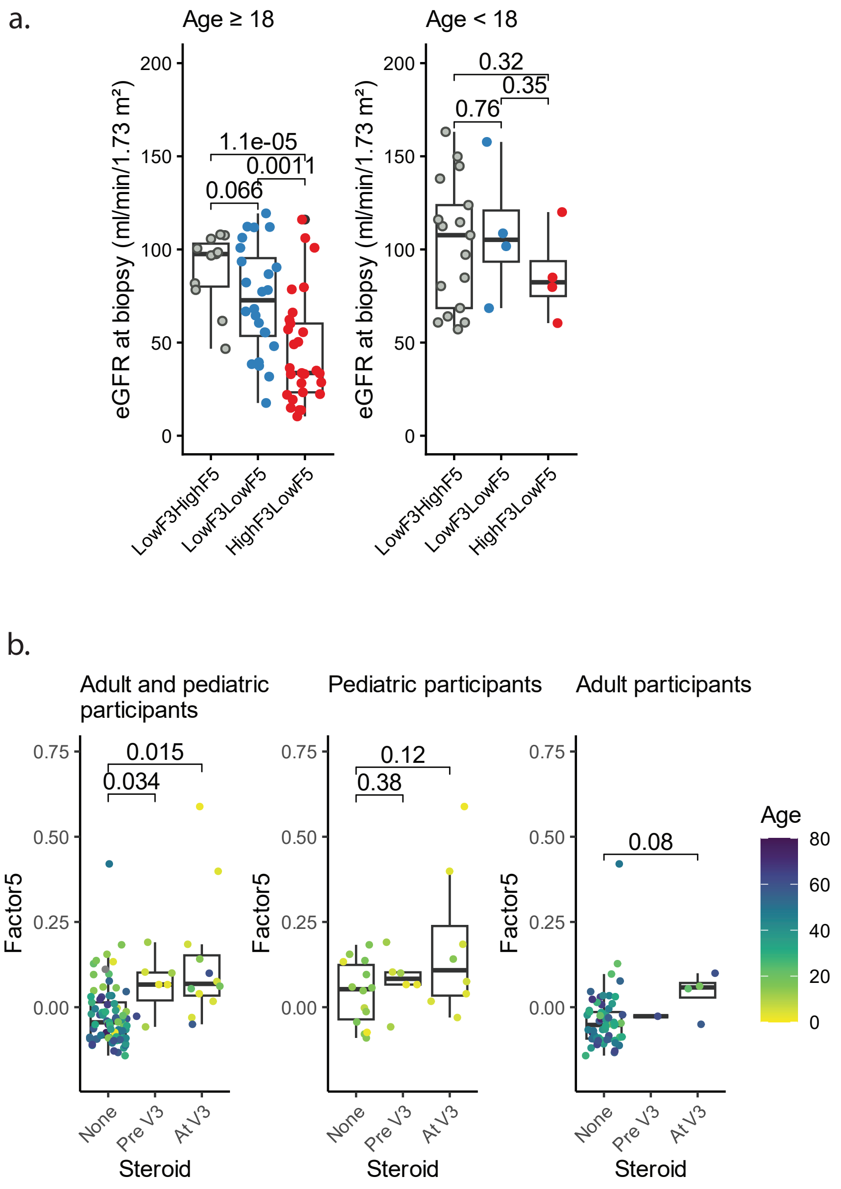


**SFig.S2. Clinical information from NEPTUNE participants and Factor3-Factor5 status.**
**a.** eGFR (ml/min/1.73 m²) across the stratified groups defined by Factor3-Factor5, constrained on adult participants (left panel) and pediatric participands (right panel). **b.** Factor 5 scores per pre-biopsy steroid exposure group across NEPTUNE for adult and pediatric participants (left panel), pediatric participants (middle panel) and adult participants (right panel). a. and b. show p-values for two-tailed t-tests.

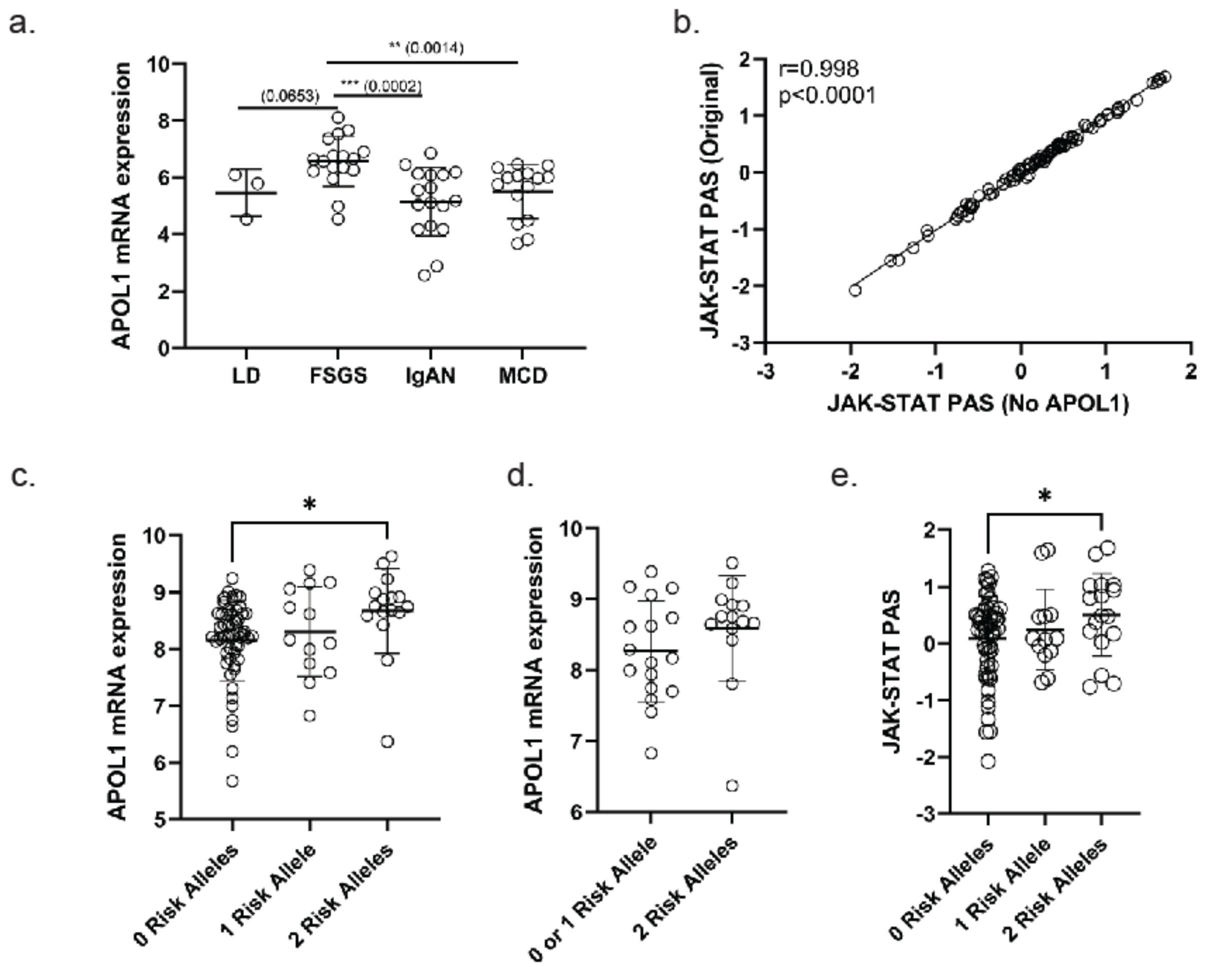


**SFigS3. JAK-STAT-APOL1 expression in podocytes of participants with FSGS. a.** Podocyte Psuedobulk of APOL1 expression based on 10x profiles **b.** JAK-STAT pathway activity score (PAS) without APOL1 in bulk glom RNAseq profiles was aligned with pathway activity using previously reported signature. **c. and d.** APOL1 expression in glomerular RNAseq profiles grouped by the number of APOL1 risk alleles in patients with FSGS and among black participants only, and **e.** JAK-STAT PAS in bulk glomerular RNAseq profiles of patient with FSGS.

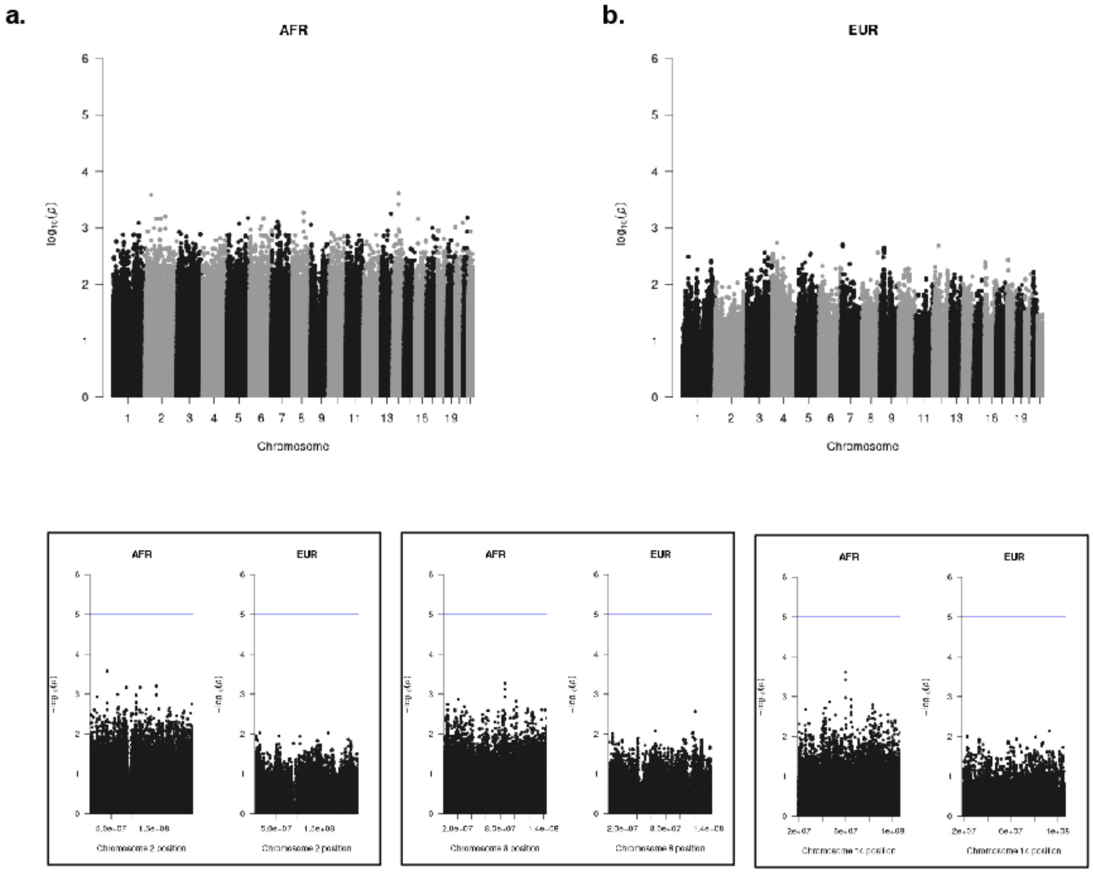


**SFigS4: Whole genome analysis of HR cases and HR controls failed to identify genetic modifiers with genome-wide significance.** Low-pass whole-genome sequencing (LP-WGS) was performed on genomic DNA from 57 African American individuals with APOL1 high-risk (HR) genotypes. After alignment to GRCh38 and variant filtering using GATK Best Practices and PLINK, ancestry-aware GWAS was conducted using Tractor, integrating local ancestry inferred by RFMix. Association results stratified by **a.** African (AFR) and **b.** European (EUR) ancestry were visualized with Manhattan plots. **c.** HR Cases (n=27) and HR Controls (n=30) identified non-coding SNPs with nominal significance in HR Cases. These SNPs did not replicate in independent cohorts of patients with AMKD.

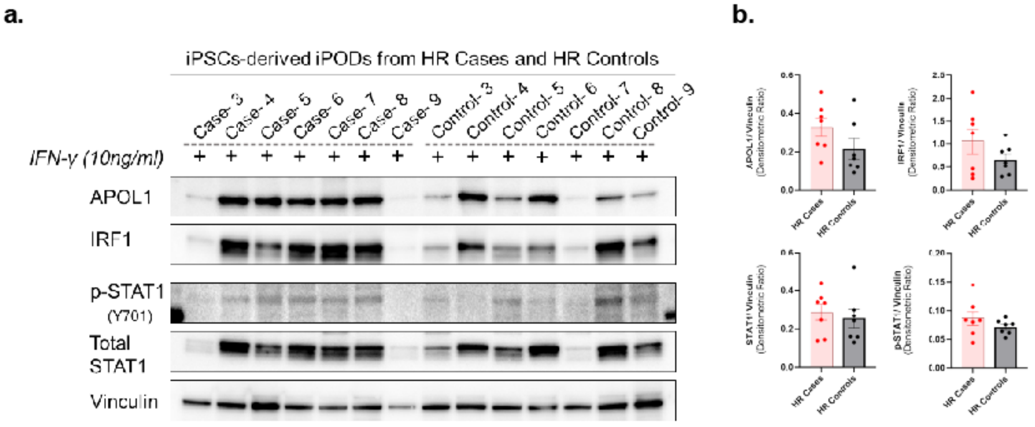


**SFigS5. IFN-γ treatment induces stronger JAK-STAT signaling in iPODs from HR Cases (n = 7) compared to HR Controls (n = 7). a.** Western blot analysis shows increased expression of key components of the JAK-STAT pathway, including APOL1, IRF1, phospho-STAT1 (Y701), and total STAT1, following 24-hour treatment with IFN-γ (10 ng/mL). In this independent second cohort of iPODs, enhanced activation was observed in 5 out of 7 HR Cases, whereas only 2 out of 7 HR Controls exhibited JAK-STAT activation.
**b.** Quantification of band intensities by densitometric analysis confirms higher pathway activation in HR Cases relative to HR Controls.
